# Healthcare System Resilience and Adaptation: A Six-Year Analysis of Heart Failure Care in the Veterans Affairs System

**DOI:** 10.1101/2025.10.13.25337954

**Authors:** Nicholas K. Brownell, Moira Inkelas, Lillian Chen, Mei Leng, Anthony Anh Quoc Doan, Nicholas Jackson, Priscilla Hsue, Michael Ong, Elizabeth Yano, Gregg C. Fonarow, Boback Ziaeian

**Author notes:** **Corresponding Author:** Boback Ziaeian, MD PhD. Division of Cardiology, David Geffen School of Medicine at University of California, Los Angeles, 650 Charles E. Young Dr. S., A2-237 CHS, Los Angeles, CA, 90095, USA.

## Abstract

**Background:** The Veterans Health Administration (VA) healthcare system cares for almost 200,000 Veterans with heart failure with reduced ejection fraction (HFrEF). It is not known how this population fared regarding continuity of care quality measures and access over the past 6 years, including the COVID-19 pandemic and its aftermath. This study aimed to evaluate temporal trends in heart failure care delivery and identify prognostic factors for clinical outcomes for Veterans with HFrEF from 1/1/2019-12/31/2024.

**Methods:** An interrupted time series analysis examined care patterns across pre-pandemic, early COVID, and late COVID periods using linear regression models with monthly fixed effects. Survival analysis of Veterans with HFrEF identified prognostic factors for mortality and hospitalization.

**Results:** The study included 210,535 individuals (mean age 73.1; 2.2% women; mean ejection fraction 32.5%). HFrEF medication prescriptions were maintained or improved, with upward trends for most medications in the early COVID period and all medications in the late COVID period (range: 0.1-0.9 percentage point increase per 6-week period, compared to pre-COVID baseline), yet with significant treatment gaps for all medication classes by study end. The onset of COVID led to a 2-4-fold increase in telehealth utilization (primary care: 18.2 percentage points, 95% CI: 14.7 to 21.6; cardiology: 8.6 percentage points, 95% CI: 7.4 to 9.8). There was a sustained 45% relative decrease in all-cause hospitalization and 40% relative decrease in heart failure hospitalization, but a sustained 20% increase in mortality. Cumulative survival was 56.1% at 6 years.

**Conclusions:** HFrEF medication rates were generally sustained or improved but well below optimal levels for Veterans with HFrEF. Telehealth replaced some in-person care early in the pandemic. Despite these changes, this high-risk population experienced a sustained decrease in hospitalizations and increase in mortality.

**Clinical Perspective:** *What is new?:* Veterans with heart failure with reduced ejection fraction increasingly used guideline directed medical therapy over the past 6 years with significant gaps remaining. There was an increased use of telehealth, sustained decrease in hospitalizations, and sustained increase in all-cause mortality for this patient population, compared to pre-COVID trends.

*What are the clinical implications?:* The sustained increase in mortality despite maintained medication access and successful telehealth implementation suggests that traditional care quality measures may not fully capture patient care associated with outcomes during major healthcare disruptions, warranting further investigation and validation in other health systems.

## Introduction

The Veterans Health Administration (VA) healthcare system is the largest integrated healthcare network in the US. Within VA, heart failure (HF) is among the top reasons for hospitalization.^1^ Guideline-directed medical therapy (GDMT) is a group of four medication classes that can dramatically improve health outcomes for persons with HF with reduced ejection fraction (HFrEF); the combination of all four medications can reduce 2-year mortality by 74%, leading to a number needed to treat of just 4.^2–4^ As a result, there is growing interest in characterizing and optimizing the care of the ∼200,000 Veterans living with HF in the contemporary era.

The coronavirus-19 (COVID-19) pandemic caused major shifts in healthcare delivery, creating a natural experiment in healthcare system resilience and adaptation. While telemedicine use increased across the nation for both primary care and subspecialties,^5–8^ the changes in care delivery patterns raise important questions about whether medication use and mortality outcomes for patients with HFrEF were simultaneously affected – either through delays in care, changes in medication delivery, or altered access patterns via telehealth. Understanding temporal changes in survival for Veterans with HFrEF is particularly important given this population may face higher risks during healthcare disruptions, whether due to advanced age, high comorbidity burden, or reliance on regular subspecialty care for optimal outcomes.^9–12^

To address these concerns, this study provides a comprehensive description of care quality for Veterans with longstanding HFrEF within the context of the COVID-19 pandemic. Through survival analysis, we identified key prognostic factors for mortality and hospitalization in Veterans with HFrEF and characterized overall survival patterns during the 6 year period. We further assessed whether the COVID-19 pandemic led to significant changes in GDMT use, telehealth services, in-person appointments, hospitalizations, and mortality for the HFrEF population at VA, and whether these changes persisted 4 years after pandemic onset.

## Methods

### Study Design and Population

We identified a retrospective population of Veterans using data derived from VA administrative claims and electronic health records. The study included Veterans diagnosed with HFrEF at VA between January 1, 2019 and December 31, 2024. A validated natural language processing algorithm identified left ventricular ejection fraction (LVEF) using echocardiography reports and clinical documentation; patients with a diagnosis of HF based on ICD code and a documented reduced LVEF (defined as ≤40%) within the past 2 years were included.^13–15^ Patients with HF with improved ejection fraction (HFimpEF) – defined as a prior reduced LVEF that recovered to >40% within the past 2 years – were included in medication and time-to-event analyses to reflect contemporary practice where these patients continue GDMT.^4^ Patients were at least 18 years old and active within the VA system, defined as having either a visit or medication fill in the past 2 years. Patients with HF with preserved/mid-range ejection fraction, history of cardiac transplantation, or left ventricular assist device were excluded. The study was approved by the VA Los Angeles IRB.

### Data Source and Variables

The VA’s central data depository, the Corporate Data Warehouse, maintains patient records including demographic data, clinical encounters, diagnoses and procedure codes, laboratory and echocardiography values, and pharmacy dispensing records. Race and ethnicity were categorized into four groups: White, Black, Hispanic, and Other (including Asian, Other, and Unknown/Declined to Answer). Geographic region was coded according to US Census Bureau definitions (Midwest, Northeast, South, West). Clinical variables included age at study initiation, sex, body mass index (BMI), heart rate, systolic and diastolic blood pressure, hemoglobin, estimated glomerular filtration rate (eGFR), and LVEF. Cardiovascular, psychiatric, and other comorbidities were identified with ICD-10 codes.

### Interrupted Time Series Analysis

To evaluate the impact of COVID-19 on the VA HFrEF population, we utilized an interrupted time series analysis. Periods were defined as pre-pandemic (1/1/2019-2/29/2020), early COVID (3/1/2020-7/31/2021), and late COVID (8/1/2021-12/31/2024). Outcomes included medication use, visit data, hospitalizations, and mortality.

VA Pharmacy data as well as non-VA medication documentation were used to establish rates of GDMT receipt for HFrEF as a 6-week rolling performance metric to smooth prescription filling variations. GDMT performance measures assessed active prescriptions for guideline-recommended medication use:^4^ angiotensin-converting enzyme inhibitor (ACEI), angiotensin receptor blocker (ARB), or angiotensin receptor-neprilysin inhibitor (ARNI); ARNI only; evidence-based beta-blocker (bisoprolol fumarate, carvedilol phosphate, or metoprolol succinate); mineralocorticoid receptor antagonist (MRA); and sodium-glucose cotransporter 2 inhibitor (SGLT2i). Patients were excluded from a medication measure if they had documented allergies to the GDMT medication being evaluated and were excluded from all measures except beta-blocker usage if they had an estimated glomerular filtration rate (eGFR) ≤60 ml/min/1.73m^2^. Rates (%) were calculated as the total number of HFrEF and HFimpEF patients on medication, divided by the total number of HFrEF patients + total number of HFimpEF patients on medication. If GDMT was prescribed and entered as a Non-VA Medication into the electronic medical record, this was also considered an active prescription.^16^

Other outcomes included monthly rates of care utilization, defined as primary care outpatient visits, cardiology outpatient visits, primary care telehealth visits, and cardiology telehealth visits. Telehealth visits included telephone, video, and Clinical Video Telehealth (CVT) visits; the latter is a specialty form of telehealth that is VA-specific.^17^ CPT codes were used to identify visit type. Clinical outcomes included monthly hospitalization rates and mortality rates. Hospitalization was further deconstructed into all-cause, HF-specific, and COVID-specific. ICD-10 codes for primary diagnosis were used to clarify the primary reason for hospitalization. Mortality was assessed by VA data as well as linkage to the National Death Index and Medicare administrative databases.

For all outcomes, interrupted time series regression models were used. Models were compared with and without monthly fixed effects to assess for seasonality. Models used robust standard errors to account for autocorrelation and heteroskedasticity. Models included linear time trends, period indicators for the three periods, and interaction terms between time and period to describe change in trends. Populations were updated every 6 weeks (medications) or every month (visits, hospitalizations, mortality) to allow for dynamic entry and exit of patients due to death, healthcare system departure, or new enrollment. All analyses were run in Stata 18 (StataCorp, College Station, TX).

### Survival Analysis

Cox proportional hazards regression identified prognostic factors for three time-to-event outcomes: all-cause mortality, first all-cause hospitalization, and a composite of death or first hospitalization. A closed cohort derived from month 1 of the prior database was defined by a start date of 1/1/2019 with follow-up until the event of interest, death, or 12/31/2024. Missing clinical variables were addressed using race-stratified multiple imputation across 5 datasets.^18,19^ Models included demographics, clinical variables, and comorbidities. Model discrimination was assessed using Harrell’s C-statistic. The proportional hazards assumption was tested using Schoenfeld residuals (global p >0.05). Complete cases were defined as patients with no missing values in key covariates and were used as a sensitivity analysis. Analyses were run in Stata. Further details can be found in **Supplemental Methods**.

## Results

The cohort included 210,535 individuals at baseline (mean age 73.1; 2.2% women; 69.5% Non-Hispanic White, 19.1% Non-Hispanic Black, 4.3% Hispanic; mean LVEF 32.5%) (**Table 1; Figure S1**). For interrupted time series analysis, March 2020 was chosen as breakpoint 1 given the declaration of a National Pandemic Emergency.^20^ F-statistic optimization using variance weighted averages of all potential breakpoints from month 22 to 48 revealed August 2021 as the superior model fit for breakpoint 2 across outcomes;^21^ this corresponded to the peak of the Delta variant and the highest COVID hospitalizations since the initial surge (**Figure S2**).^22^ Except for medication rates, seasonally adjusted models showed significantly better fit across outcomes and were presented (**Table S1**).

**Table 1:** Demographics and Comorbidities for Patients at Baseline.

| **Characteristic** | **Statistic** |
| --- | --- |
| **Sample Size** | N = 210,535 |
| **DEMOGRAPHICS** |  |
| **Age, years** | 73.1 (10.6) |
| **Female sex** | 4,639 (2.2%) |
| **Race/Ethnicity** |  |
| **White** | 146,221 (69.5%) |
| **Black** | 40,172 (19.1%) |
| **Hispanic** | 9,143 (4.3%) |
| **Other/Missing** | 14,999 (7.1%) |
| **Geographic Region** |  |
| **Midwest** | 50,328 (23.9%) |
| **Northeast** | 27,229 (12.9%) |
| **South** | 91,272 (43.4%) |
| **West** | 39,121 (18.6%) |
| **CLINICAL PARAMETERS** |  |
| **BMI (per kg/m<sup>2</sup>)** | 29.8 (6.7) |
| **Pulse (per bpm)** | 75 (14) |
| **Systolic BP (per mmHg)** | 127 (19) |
| **Diastolic BP (per mmHg)** | 72 (12) |
| **Hemoglobin (per g/dL)** | 13.0 (2.2) |
| **eGFR, mL/min/1.73m<sup>2</sup>** | 63.0 (27.9) |
| **Ejection Fraction, %** | 32.5 (14.5) |
| **COMORBIDITIES** |  |
| **Atrial fibrillation** | 91,798 (43.6%) |
| **Coronary artery disease** | 151,031 (71.7%) |
| **Chronic kidney disease** | 111,959 (53.2%) |
| **COPD** | 18,066 (8.6%) |
| **Diabetes mellitus** | 129,413 (61.5%) |
| **Hyperlipidemia** | 161,487 (76.7%) |
| **Hypertension** | 187,892 (89.3%) |
| **Cerebrovascular accident** | 31,060 (14.8%) |
| **Alcohol use disorder** | 25,552 (12.1%) |
| **Substance use disorder** | 18,327 (8.7%) |
| **Smoking** | 39,737 (18.9%) |
| **Depression** | 54,814 (26.0%) |
| **Peripheral artery disease** | 107,009 (50.1%) |
| **Cancer** | 37,158 (17.6%) |

BMI, body mass index; BP, blood pressure; COPD, chronic obstructive pulmonary disease; eGFR, estimated glomerular filtration rate.
Continuous variables presented as mean (SD); binary variables presented as N (%).

### Medication Use

In the pre-COVID year, medication rates were highest for ACE ARB ARNI (mean prescription rate: 61.56%, SD: 0.15) and beta-blocker (64.41%, SD: 0.16) (**Figure 1**). More recently approved medical therapies had the lowest baseline rates (SGLT2i: 3.98%, SD: 0.90; ARNI: 6.86%, SD: 1.21). Pre-COVID trends suggested these newer medications experienced the fastest growth in prescription rates, with ARNI growing 0.40 percentage points per 6-weeks (95% CI: 0.38 to 0.42) and SGLT2i growing 0.29 percentage points per 6-weeks (95% CI: 0.26 to 0.33). In contrast, ACE ARB ARNI and beta-blockers were relatively stable in the pre-COVID period (0.01 percentage points per 6-weeks, 95% CI: -0.02 to 0.04; 0.04 percentage points per 6-weeks, 95% CI: 0.02 to 0.06, respectively).

**Figure 1:**
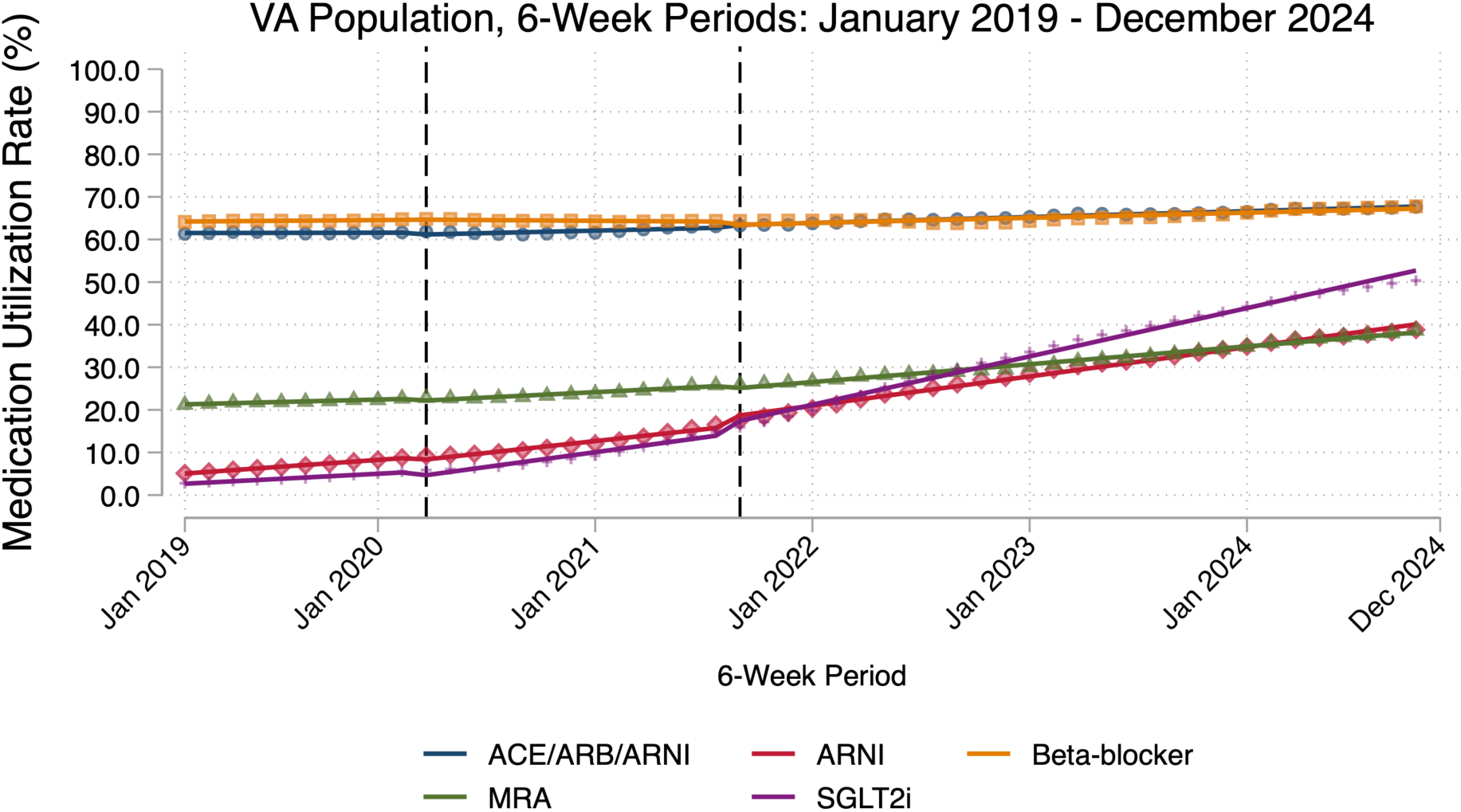
Heart Failure Medication Utilization Trends Between 2019 and 2024 ACEI: angiotensin-converting enzyme inhibitor. ARB: angiotensin receptor blocker. ARNI: angiotensin receptor-neprilysin inhibitor. MRA: mineralocorticoid receptor antagonist. SGLT2i: sodium glucose cotransporter 2 inhibitor. First vertical line indicates transition from pre-COVID to early COVID phase (March 2020). Second vertical line indicates transition from early COVID to late COVID phase (August 2021).

The onset of the pandemic in March 2020 was associated with a small but significant decrease for some medications (SGLT2i: -1.39 percentage points, 95% CI: -2.54 to -0.25; ARNI: -0.91 percentage points, 95% CI: -1.73 to -0.10; MRA: -0.64 percentage points, 95% CI: -1.19 to -0.09) (**Table 2**). Beta-blocker and ACE ARB ARNI remained constant compared to pre-COVID levels. Subsequently, there was variable growth for all medications in the early COVID period through August 2021 (range: 0.12-0.48 percentage points per 6-weeks) with the exception of beta-blockers, which saw a decrease (-0.08 percentage points per 6-weeks, 95% CI: -0.12 to -0.04), compared to pre-COVID trends.

**Table 2:**
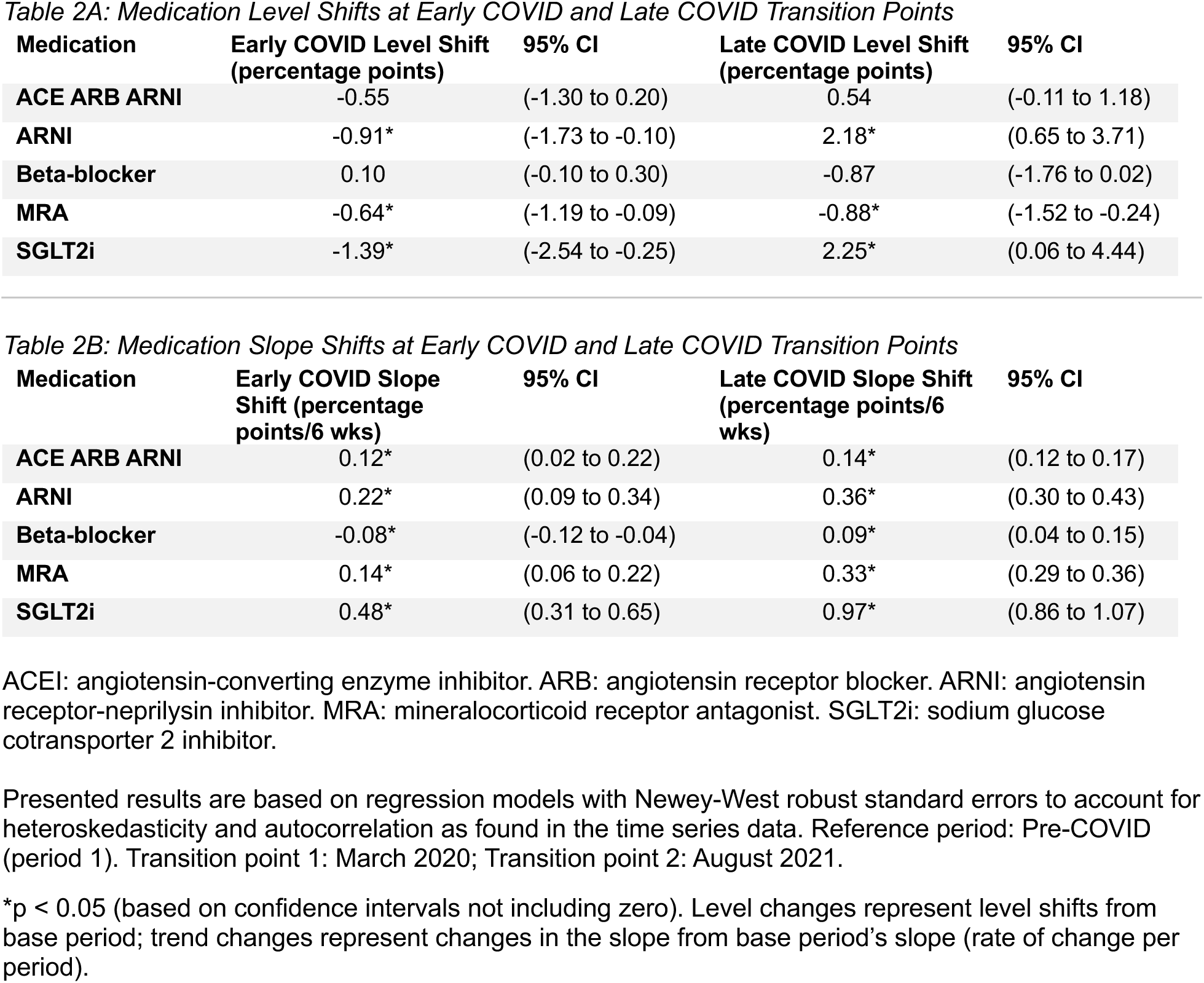
Medication Rates: Level Shifts and Slope Shifts During the Early and Late COVID Periods.

In contrast, the late pandemic period was associated with a significant increase in GDMT receipt for SGLT2i (2.25 percentage points, 95% CI: 0.06 to 4.44) and ARNI (2.18 percentage points, 95% CI: 0.65 to 3.71) but a decrease in MRA (-0.88 percentage points, 95% CI: -1.52 to -0.24), compared to pre-COVID levels. ACE ARB ARNI and beta-blocker remained stable. There was again an upward trend for all medications in the late COVID period, compared to pre-COVID trends (range: 0.09-0.97 percentage points per 6-weeks); by study end, all medication rates were higher than baseline but still below target (Range: 38-68%). Raw regression output and individual medication figures are available (**Table S2; Figure S3**).

### Visit Data

In the pre-COVID period, 23.40% (SD: 1.28%) of Veterans with HFrEF had a primary care visit each month (**Figure 2A**), most of which were via telehealth. Overall, 8.32% (SD: 0.57%) of Veterans with HFrEF had face-to-face primary care visits each month in the pre-COVID period. In contrast, 6.49% (SD: 0.49%) of Veterans with HFrEF had appointments with cardiology each month (**Figure 2B**); 4.76% (SD: 0.41%) of Veterans with HFrEF had face-to-face cardiology visits each month. Less than 2% of Veterans had telehealth cardiology appointments in the pre-COVID period. Pre-COVID trends suggested a stable healthcare system; all visit types across both primary care and cardiology had level slopes.

**Figure 2:**
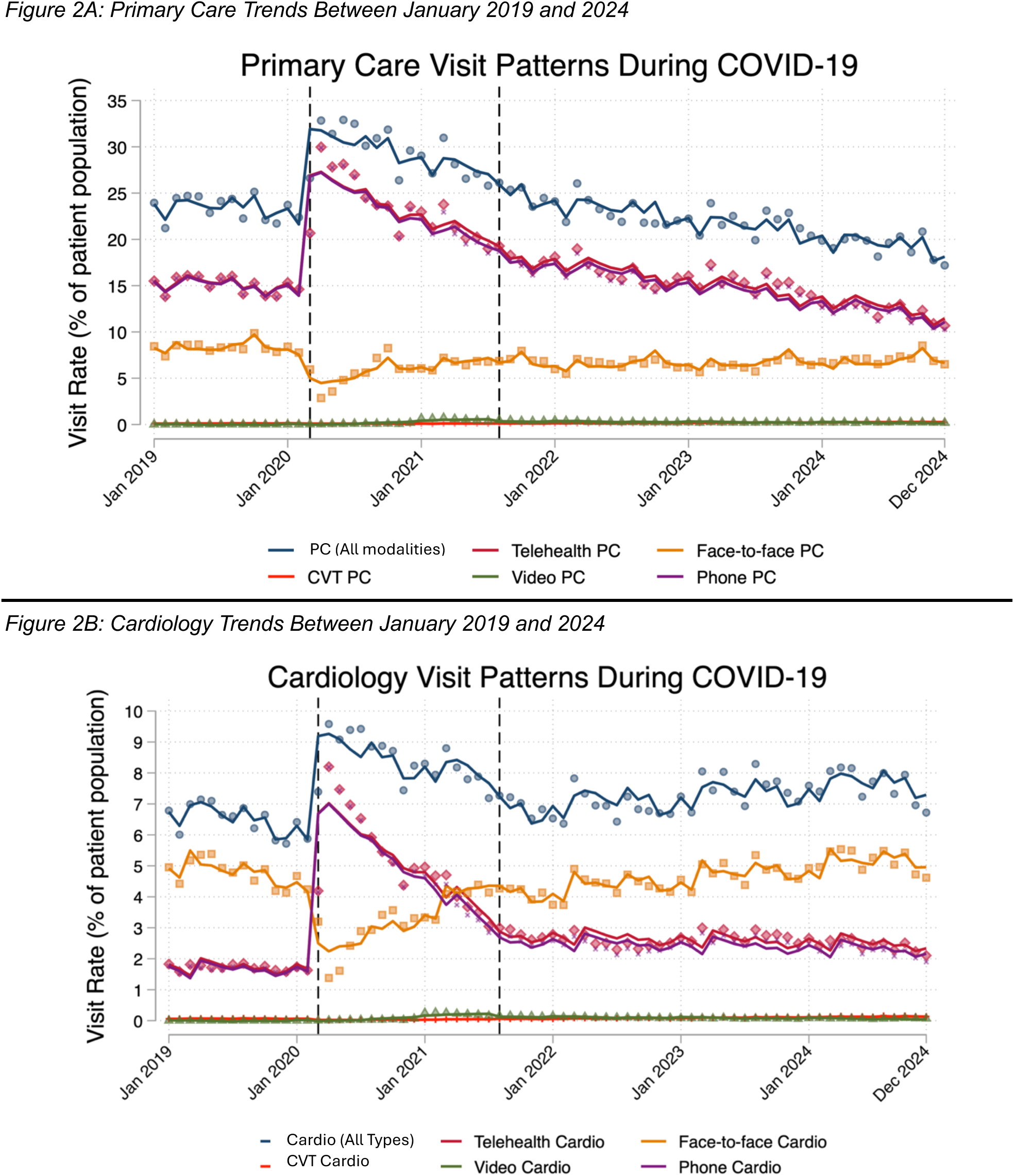
Primary Care and Cardiology Visit Trends Between 2019 and 2024 First vertical line indicates transition from pre-COVID to early COVID phase (March 2020). Second vertical line indicates transition from early COVID to late COVID phase (August 2021).

The onset of COVID yielded a drop in face-to-face visits and a rise in telehealth visits for both primary care and cardiology. Specifically, COVID led to an overall rise of 11.56 percentage points (95% CI: 8.27 to 14.85) in primary care visits, attributed to a doubling of telehealth visits (18.17 percentage points; 95% CI: 14.74 to 21.60), compared to baseline levels (**Table 3**). Face-to-face visits dropped by 6.61 percentage points (95% CI: -7.93 to -5.29), compared to baseline. Subsequently, there was a slow downward trend in telehealth visits (-0.42 percentage points per month, 95% CI: -0.64 to -0.19). Face-to-face visits saw a slow recovery in the early COVID period (0.19 percentage points per month; 95% CI: 0.11 to 0.28), compared to pre-COVID trends. Findings were similar for cardiology, with an overall rise of 3.18 percentage points (95% CI: 2.09 to 4.27) in cardiology visits at pandemic onset, attributed to a 5-fold increase in telehealth (8.62 percentage points; 7.40 to 9.84), compared to baseline levels. Face-to-face visits dropped by 5.44 percentage points (95% CI: -6.18 to -4.70). Subsequently, both telehealth and face-to-face visits trended towards pre-COVID levels.

**Table 3:**
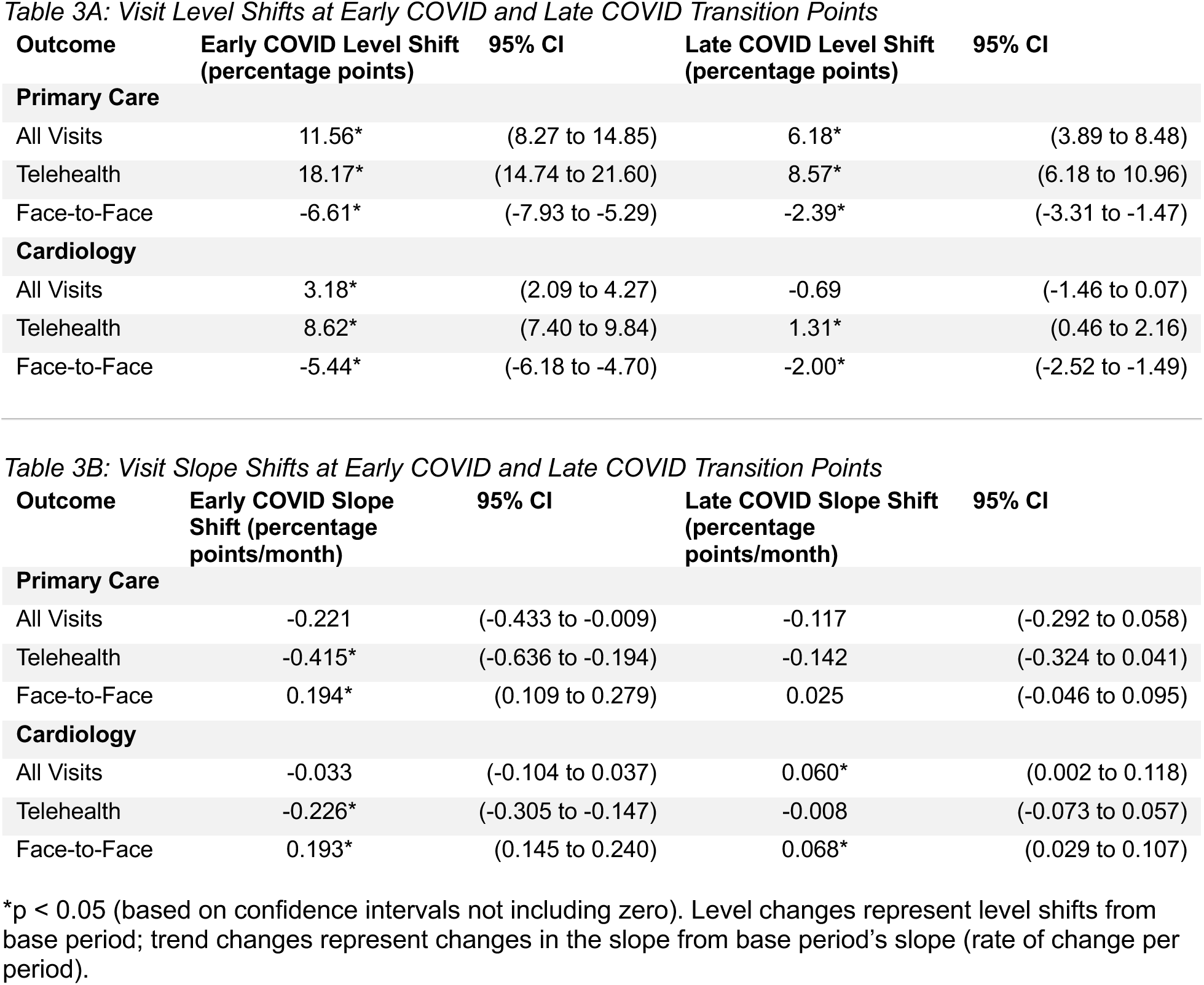
Visit Rates: Level Shifts and Slope Shifts During the Early and Late COVID Periods.

The onset of the late COVID period led to a new baseline for the modality of outpatient healthcare delivery. Primary care visits noted a significant increase of 6.18 percentage points (95% CI: 3.89 to 8.48) from baseline levels, again predominantly composed of a change in telehealth visits (8.57 percentage points, 95% CI: 6.18 to 10.96). Face-to-face visits remained below baseline, with a significant decrease of 2.39 percentage points (95% CI: -3.31 to -1.47), compared to baseline levels. Subsequently, there was no significant further recovery of primary care visits; by study end, face-to-face primary care appointment rates were slightly below baseline, with telehealth appointment rates returning to baseline.

In contrast, with the onset of the late COVID period, cardiology visits were unchanged from pre-COVID levels. However, there was an increase in telehealth (1.31 percentage points, 95% CI: 0.46 to 2.16) with a decrease in face-to-face visits (-2.00 percentage points, 95% CI: -2.52 to -1.49). Subsequently, there was again recovery in cardiology visits, compared to pre-COVID trends, at an increase of 0.06 percentage points per month (95% CI: 0.002 to 0.12). This was most notable in face-to-face visits with an increase of 0.07 percentage points per month (95% CI: 0.03 to 0.11). There was no significant change in telehealth visits in the late COVID period, compared to pre-COVID trends. Thus, by study end, there was a slight increase in appointments rates for cardiology, with face-to-face appointment rates returning to baseline, and telehealth appointment rates slightly above baseline. Raw regression output is available (**Tables S3 and S4**). Seasonal effects by month were notable in primary care face-to-face appointments (**Figure S4A**) and cardiology visits (**Figure S4B**).

### Hospitalizations and Mortality

In the pre-COVID year, the mean monthly all-cause hospitalization rate for Veterans with HFrEF was 3.81% (SD: 0.18) (**Figure 3**). Monthly HF hospitalization rate was 0.75% (SD: 0.12) and monthly mortality rate was 1.33% (SD 0.11). Pre-COVID trends suggested a stable healthcare system; all-cause hospitalization, HF hospitalization, and mortality had level slopes.

**Figure 3:**
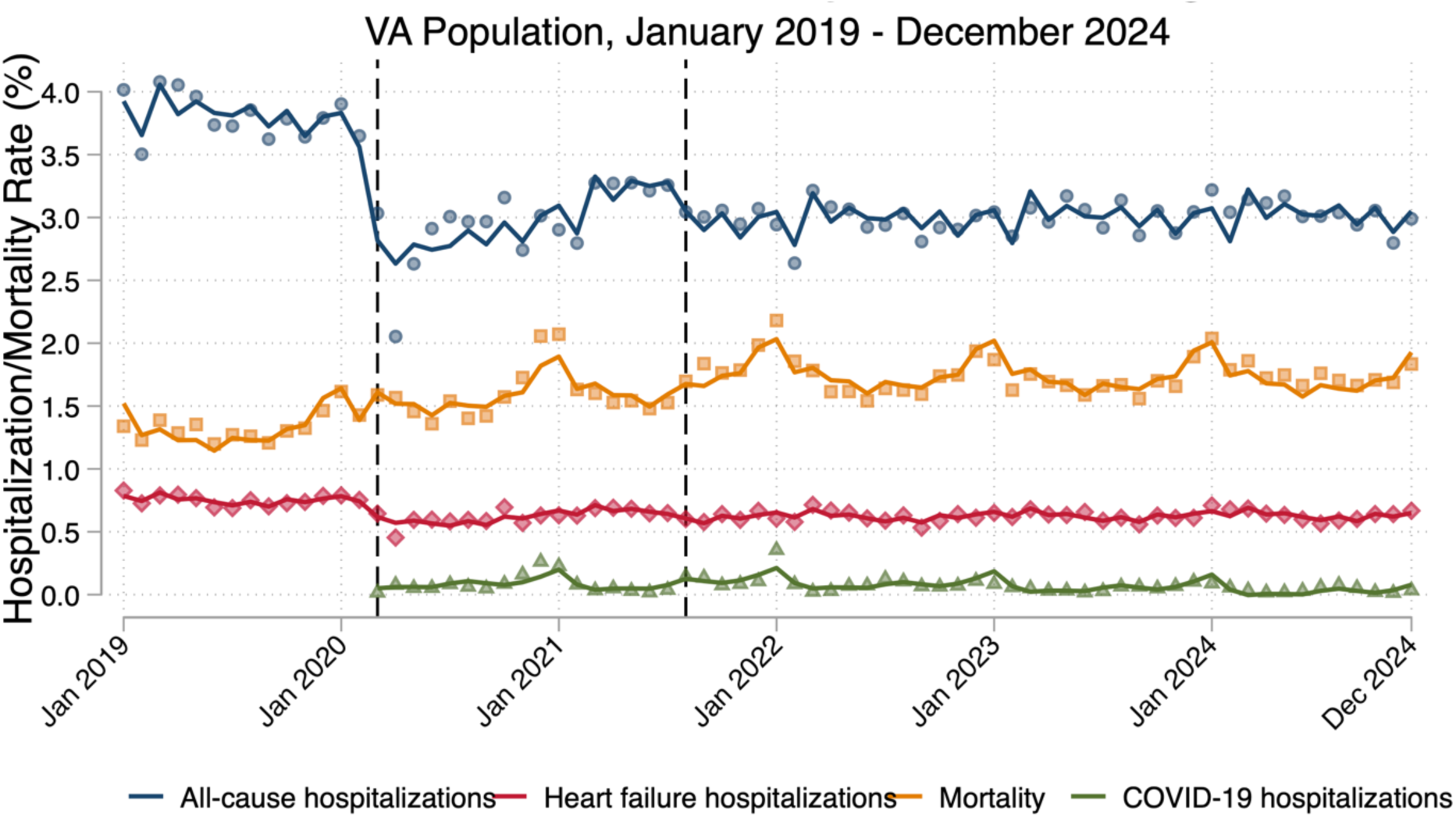
Hospitalization and Mortality Trends Between 2019 and 2024 First vertical line indicates transition from pre-COVID to early COVID phase (March 2020). Second vertical line indicates transition from early COVID to late COVID phase (August 2021).

The onset of the pandemic was associated with a significant decrease in monthly all-cause hospitalization rate by 1.90 percentage points (95% CI: -2.41 to -1.38) and a significant decrease in monthly HF hospitalization rate by 0.31 percentage points (95% CI: -0.41 to -0.22), compared to pre-COVID levels (**Table 4**); this translates to ∼45% relative decrease in all-cause hospitalizations and a ∼40% relative decrease in HF hospitalizations. Subsequently, there was an upward trend in both all-cause hospitalization rate (0.05 percentage points per month; 95% CI: 0.03 to 0.07) and HF hospitalization rate (0.008 percentage points per month; 95% CI: 0.004 to 0.012), compared to pre-COVID trends. In contrast, there was a significant increase in monthly mortality rate of 0.24 percentage points (95% CI: 0.02 to 0.45) with the onset of COVID, translating to ∼20% relative increase from pre-COVID levels. Mortality trend was stable in the early COVID period, compared to the pre-COVID trends.

**Table 4:**
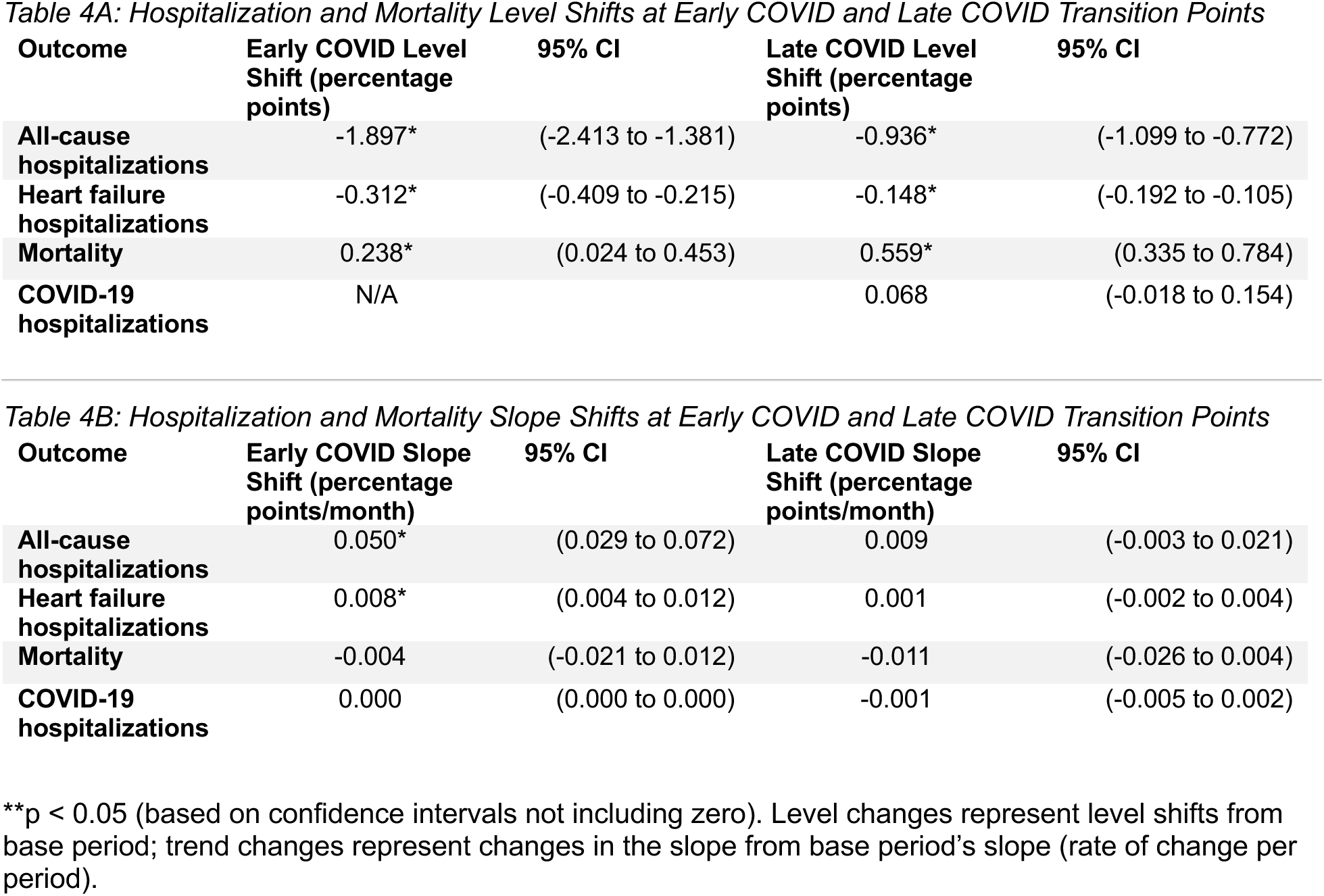
Hospitalization and Mortality: Level Shifts and Slope Shifts During the Early and Late COVID Periods.

The late COVID period was again associated with a significant but smaller decrease in monthly all-cause hospitalization rate by 0.94 percentage points (95% CI: -1.10 to -0.77) and a significant but smaller decrease in monthly HF hospitalization rate by 0.15 percentage points (95% CI: -0.19 to -0.11), compared to pre-COVID levels. There was another significant increase in monthly mortality rate of 0.56 percentage points (95% CI: 0.34 to 0.78), translating to a ∼40% increase from pre-COVID levels. Hospitalization trends, including for COVID, were stable compared to pre-COVID trends; for all-cause hospitalization and HF hospitalization, this suggested a new baseline lower than the pre-COVID period. Similarly, mortality in the late COVID period was stable compared to the pre-COVID period, suggesting an ongoing trend of roughly 500 additional deaths per 100,000 patients each month, compared to the pre-COVID mortality rate. Raw regression output and individual outcome figures are available (**Table S5; Figure S5**). Seasonal effects were most significant for mortality and COVID-19 hospitalizations, where summer months noted the lowest mortality rates, followed by spring and fall, followed by winter (**Figure S6**).

### Survival Analysis

A total of 210,535 Veterans were included at baseline; complete case analysis included 139,169 Veterans (**Table S6**). Race distribution was maintained across imputed datasets. There was high prevalence of cardiac and noncardiac comorbidities, including coronary artery disease (71.7%), hypertension (89.3%), and hyperlipidemia (76.7%). Kaplan-Meier survival curves demonstrated overall survival in the cohort over 6 years of follow-up, with cumulative survival of 90.5% at 1 year, 81.6% at 3 years, and 56.1% at 6 years; cumulative hospitalization-free survival of 85.0% at 1 year, 79.2% at 3 years, and 66.7% at 6 years; and cumulative probabilities of being free from the composite endpoint of 78.9% at 1 year, 67.8% at 3 years, and 43.2% at 6 years (**Figure S7**).

After multiple imputation, several variables showed time-varying effects based on proportions hazard violations (p <0.001), including age, BMI, blood pressure, heart rate, atrial fibrillation, CKD, COPD, depression, cancer, hemoglobin, and LVEF; single hazard ratio estimates are thus best interpreted as average effects over the study period. The multiple imputation analysis revealed distinct predictors of mortality, hospitalization, and the composite outcome (**Table 5**). Age demonstrated opposing effects, increasing mortality risk by 2.5% per year (HR 1.025, 95% CI 1.024 to 1.026) while reducing all-cause hospitalization risk by 1.1% per year (HR 0.989, 95% CI 0.989 to 0.990). Female sex was protective across all outcomes, reducing risk by 10-20% (mortality HR 0.797, 95% CI 0.757 to 0.839; hospitalization HR 0.906, 95% CI: 0.857 to 0.958; composite HR 0.871, 95% CI: 0.835 to 0.909). Clinical parameters noted a higher BMI was associated with reduced mortality (HR 0.991 per kg/m², 95% CI: 0.989 to 0.992) and composite risk (0.996 per kg/m², 95% CI: 0.995 to 0.998) while showing no association with all-cause hospitalization. Each incremental increase in LVEF was associated with a reduction in risk for all outcomes (mortality HR 0.997 (95% CI: 0.996 to 0.997), hospitalization HR 0.985 (95% CI: 0.984 to 0.985), composite HR 0.993 (95% CI: 0.992 to 0.993)). Comorbidity burden generally increased risk universally, with substance use disorder and smoking noting the strongest associations across outcomes. Hypertension and hyperlipidemia had higher associations with hospitalization than mortality. Model discrimination was modest (C statistics 0.584-0.624) reflecting the complex interplay of risk factors in this VA population. Findings were robust to missing data approach with complete cases only (**Table S7**).

**Table 5:**
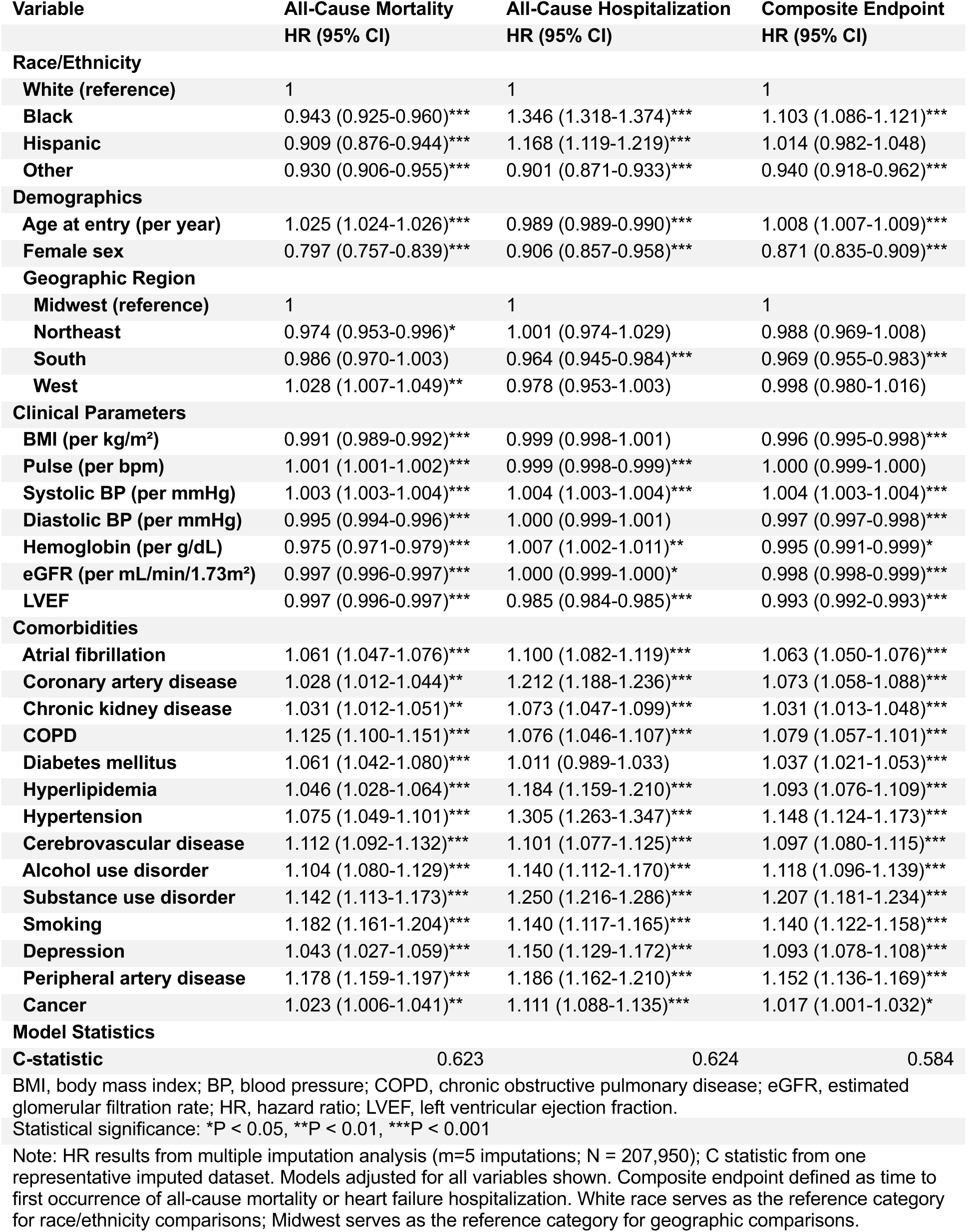
Cox Proportional Hazard Models for Mortality and Hospitalization.

| Variable | All-Cause Mortality<br>HR (95% CI) | All-Cause Hospitalization<br>HR (95% CI) | Composite Endpoint<br>HR (95% CI) |
| --- | --- | --- | --- |
| **Race/Ethnicity** |  |  |  |
| White (reference) | 1 | 1 | 1 |
| Black | 0.943 (0.925-0.960)*** | 1.346 (1.318-1.374)*** | 1.103 (1.086-1.121)*** |
| Hispanic | 0.909 (0.876-0.944)*** | 1.168 (1.119-1.219)*** | 1.014 (0.982-1.048) |
| Other | 0.930 (0.906-0.955)*** | 0.901 (0.871-0.933)*** | 0.940 (0.918-0.962)*** |
| **Demographics** |  |  |  |
| Age at entry (per year) | 1.025 (1.024-1.026)*** | 0.989 (0.989-0.990)*** | 1.008 (1.007-1.009)*** |
| Female sex | 0.797 (0.757-0.839)*** | 0.906 (0.857-0.958)*** | 0.871 (0.835-0.909)*** |
| **Geographic Region** |  |  |  |
| Midwest (reference) | 1 | 1 | 1 |
| Northeast | 0.974 (0.953-0.996)* | 1.001 (0.974-1.029) | 0.988 (0.969-1.008) |
| South | 0.986 (0.970-1.003) | 0.964 (0.945-0.984)*** | 0.969 (0.955-0.983)*** |
| West | 1.028 (1.007-1.049)** | 0.978 (0.953-1.003) | 0.998 (0.980-1.016) |
| **Clinical Parameters** |  |  |  |
| BMI (per kg/m<sup>2</sup>) | 0.991 (0.989-0.992)*** | 0.999 (0.998-1.001) | 0.996 (0.995-0.998)*** |
| Pulse (per bpm) | 1.001 (1.001-1.002)*** | 0.999 (0.998-0.999)*** | 1.000 (0.999-1.000) |
| Systolic BP (per mmHg) | 1.003 (1.003-1.004)*** | 1.004 (1.003-1.004)*** | 1.004 (1.003-1.004)*** |
| Diastolic BP (per mmHg) | 0.995 (0.994-0.996)*** | 1.000 (0.999-1.001) | 0.997 (0.997-0.998)*** |
| Hemoglobin (per g/dL) | 0.975 (0.971-0.979)*** | 1.007 (1.002-1.011)** | 0.995 (0.991-0.999)* |
| eGFR (per mL/min/1.73m<sup>2</sup>) | 0.997 (0.996-0.997)*** | 1.000 (0.999-1.000)* | 0.998 (0.998-0.999)*** |
| LVEF | 0.997 (0.996-0.997)*** | 0.985 (0.984-0.985)*** | 0.993 (0.992-0.993)*** |
| **Comorbidities** |  |  |  |
| Atrial fibrillation | 1.061 (1.047-1.076)*** | 1.100 (1.082-1.119)*** | 1.063 (1.050-1.076)*** |
| Coronary artery disease | 1.028 (1.012-1.044)** | 1.212 (1.188-1.236)*** | 1.073 (1.058-1.088)*** |
| Chronic kidney disease | 1.031 (1.012-1.051)** | 1.073 (1.047-1.099)*** | 1.031 (1.013-1.048)*** |
| COPD | 1.125 (1.100-1.151)*** | 1.076 (1.046-1.107)*** | 1.079 (1.057-1.101)*** |
| Diabetes mellitus | 1.061 (1.042-1.080)*** | 1.011 (0.989-1.033) | 1.037 (1.021-1.053)*** |
| Hyperlipidemia | 1.046 (1.028-1.064)*** | 1.184 (1.159-1.210)*** | 1.093 (1.076-1.109)*** |
| Hypertension | 1.075 (1.049-1.101)*** | 1.305 (1.263-1.347)*** | 1.148 (1.124-1.173)*** |
| Cerebrovascular disease | 1.112 (1.092-1.132)*** | 1.101 (1.077-1.125)*** | 1.097 (1.080-1.115)*** |
| Alcohol use disorder | 1.104 (1.080-1.129)*** | 1.140 (1.112-1.170)*** | 1.118 (1.096-1.139)*** |
| Substance use disorder | 1.142 (1.113-1.173)*** | 1.250 (1.216-1.286)*** | 1.207 (1.181-1.234)*** |
| Smoking | 1.182 (1.161-1.204)*** | 1.140 (1.117-1.165)*** | 1.140 (1.122-1.158)*** |
| Depression | 1.043 (1.027-1.059)*** | 1.150 (1.129-1.172)*** | 1.093 (1.078-1.108)*** |
| Peripheral artery disease | 1.178 (1.159-1.197)*** | 1.186 (1.162-1.210)*** | 1.152 (1.136-1.169)*** |
| Cancer | 1.023 (1.006-1.041)** | 1.111 (1.088-1.135)*** | 1.017 (1.001-1.032)* |
| **Model Statistics** |  |  |  |
| C-statistic | 0.623 | 0.624 | 0.584 |

BMI, body mass index; BP, blood pressure; COPD, chronic obstructive pulmonary disease; eGFR, estimated glomerular filtration rate; HR, hazard ratio; LVEF, left ventricular ejection fraction.
Statistical significance: \*P < 0.05, \*\*P < 0.01, \*\*\*P < 0.001
Note: HR results from multiple imputation analysis (m=5 imputations; N = 207,950); C statistic from one representative imputed dataset. Models adjusted for all variables shown. Composite endpoint defined as time to first occurrence of all-cause mortality or heart failure hospitalization. White race serves as the reference category for race/ethnicity comparisons; Midwest serves as the reference category for geographic comparisons.

## Discussion

Our findings characterize the past 6 years of HF care within the VA healthcare system. Medication rates were either sustained or improved throughout the study. The existing VA telehealth model increased in use through the peak of the pandemic. Hospitalizations declined and sustained rates at a lower level than the pre-pandemic period; in contrast, mortality increased and has sustained rates at a higher level. In survival analysis, age, male sex, and numerous comorbidities were associated with increased mortality over time.

Extrapolating from pre-COVID trends, the onset of COVID in March 2020 potentially limited 500-1200 Veterans from receiving GDMT, with newer medications like SGLT2i and ARNI most impacted (**Table S8**). This amounted to only ∼1% of the eligible HF population, and all medications recovered impressively, with observed prescriptions at study end being between 400 and 19,000 higher than expected based on pre-COVID trends. Again, newer therapies like SGLT2i and ARNI noted the most accelerated adoption during the early and late COVID periods. Most importantly, no medications had observed rates that fell relative to pre-COVID trends. In sum, most Veterans with HFrEF received GDMT throughout the pandemic and afterwards. However, these rates for each class of medication remained well below ideal rates and those reported for HF specialty programs.^2^

These findings are supported by similar studies from the early COVID period in Veterans with recently diagnosed HFrEF, whose authors suggest that telehealth may have provided continued access through the pandemic.^23^ Outside of VA, a study of adults with chronic diseases – including HF – revealed COVID-19-related medication barriers were common, with 51% of respondents reporting a medication-related problem attributed to COVID’s disruption of the healthcare system, and ∼20% noting difficulty obtaining their medications because of offices closed for in-person visits.^24^ A systematic review noted that COVID led to major interruptions in chronic disease management, particularly related to difficulty in reaching physicians and receiving medications in a timely manner.^25^ The VA integrated healthcare system model appeared robust, in particular its pharmacy infrastructure, which maintained medication access despite massive healthcare disruptions.

In contrast to settings outside VA, the VA was already delivering a considerable amount of telehealth prior to the pandemic and expanded considerably.^17^ Both primary care and cardiology saw a rapid and meaningful growth in telehealth use before partially returning to traditional face-to-face visits in the late COVID period. This pattern aligns with other specialties at VA, other healthcare systems, and across insurance coverage options.^26–29^ The rapid transition to telehealth was thus not a unique solution for the VA; a systematic review noted multiple studies reporting telehealth as the primary solution to minimize treatment discontinuation and ensure high adherence.^25^ Other healthcare systems also noted similar medication rates between HF patients with telehealth visits and those with in-person visits immediately after the pandemic.^30^ Importantly, in studies both within and outside VA, transition to telehealth was not associated with worsening of clinical outcomes.^5,31,32^ Altogether, this suggests some degree of telehealth adoption as the new state of US healthcare delivery.

A marked finding is a reduction in both all-cause and HF hospitalizations but increased mortality for the Veteran HFrEF population that sustained into the late COVID period. While lower volume of admissions for HF patients is documented in the early pandemic,^33^ broader national trends suggest improving age-adjusted mortality rates for cardiovascular disease since the peak of the pandemic.^34^ The elevated mortality in our study, despite maintained medication rates, successful transition to telehealth, and reduced hospitalizations persistent over several years, may reflect a “new normal.” Such findings reveal important limitations in traditional healthcare quality metrics during system-wide disruptions; process measures may be insufficient indicators of care quality in settings of massive healthcare system disruption when patients face barriers beyond the healthcare system’s direct control, such as delayed care-seeking, social isolation, financial hardship, or deferred preventive care. The persistence of the mortality finding four years post-pandemic onset may indicate a long-lasting shift in baseline risk for this highly vulnerable population or may relate to unmeasured confounders; further monitoring is clearly vital and warranted.

Our observed 6-year survival rate of 56.1% for Veterans with HFrEF is largely consistent with other national HF data, and demographic predictors were similarly consistent with the literature.^4^ Female sex was associated with better survival;^35–37^ however, female sex was also associated with lower hospitalization risk, which contrasts with prior studies noting higher hospitalization rates among women with HF.^36,38,39^ Otherwise traditional prognostic factors including lower hemoglobin and LVEF, as well as comorbidities like smoking and substance use, maintained their expected association with worse outcomes.^4,40–42^

### Limitations

There are important limitations to address. First, outpatient visits and hospitalizations outside of the VA system were not included in the present analysis and thus our study does not account for the recent rise in community care for US Veterans.^43^ Second, HFimpEF is an important diagnosis that reflects recovery of LVEF; by including this subgroup in our study, we likely inflated medication rates relative to a HFrEF-only cohort. Similarly, including HFimpEF in our survival cohort may bias results to more favorable outcomes as HFimpEF patients have better survival and lower hospitalization rates compared to those with persistent HFrEF.^42,44^ Third, telehealth visits do not have a precise definition as telephone visits – which accounted for most telehealth visits – represent various types of patient contact from the primary care or cardiology clinic. It is also unknown which primary diagnoses drove changes in all-cause hospitalizations. Fourth, the study does not include medication dosing and measures of adherence. Fifth, the longitudinal analysis with a dynamic Veteran population, as well as the time-to-event analyses, aggregate all VA centers into a single analysis; future disaggregation may provide insights into secular trends and potentially regional variation that contributed to the findings.

### Conclusions

The HF population at VA largely maintained or increased use of GDMT medications throughout the pandemic and its aftermath, with a robust adaptation of telehealth during the peak of the pandemic. Notably, decrease in hospitalizations and an increase in mortality has continued into present day.

## Data Availability

The data underlying this study are protected by federal privacy regulations and cannot be shared publicly. Data may be available from the U.S. Department of Veterans Affairs for researchers who meet the criteria for access to confidential data.

## Acknowledgements

Dr. Yano’s effort was funded by a VA Health Systems Research Senior Research Career Scientist Award (RCS 05-195).

## Supplemental Material

## Supplemental Methods

For the survival analysis, the most recent clinical variable values in the year prior to 1/1/2019 were used. Race-stratified multiple imputation using chained equations across 5 datasets was performed to address missing data in clinical variables (BMI, heart rate, blood pressure, hemoglobin, eGFR, and LVEF), consistent with established guidance for large samples.^18,19^ Nelson-Aalen cumulative hazard estimators were included in the imputation model to maintain survival structure and ensure missing variables were imputed conditional on observed survival patterns within each race/ethnicity.^45^ Imputation adequacy was assessed by consistency of variable distributions across imputed datasets within each racial stratum as well as confirmation of clinical plausibility of all imputed values. Complete cases were defined as patients with no missing values in key covariates required for Cox regression models after creation of Nelson-Aalen cumulative hazard estimators. Kaplan-Meier survival curves were generated for visualization.

---

**Table S1:**
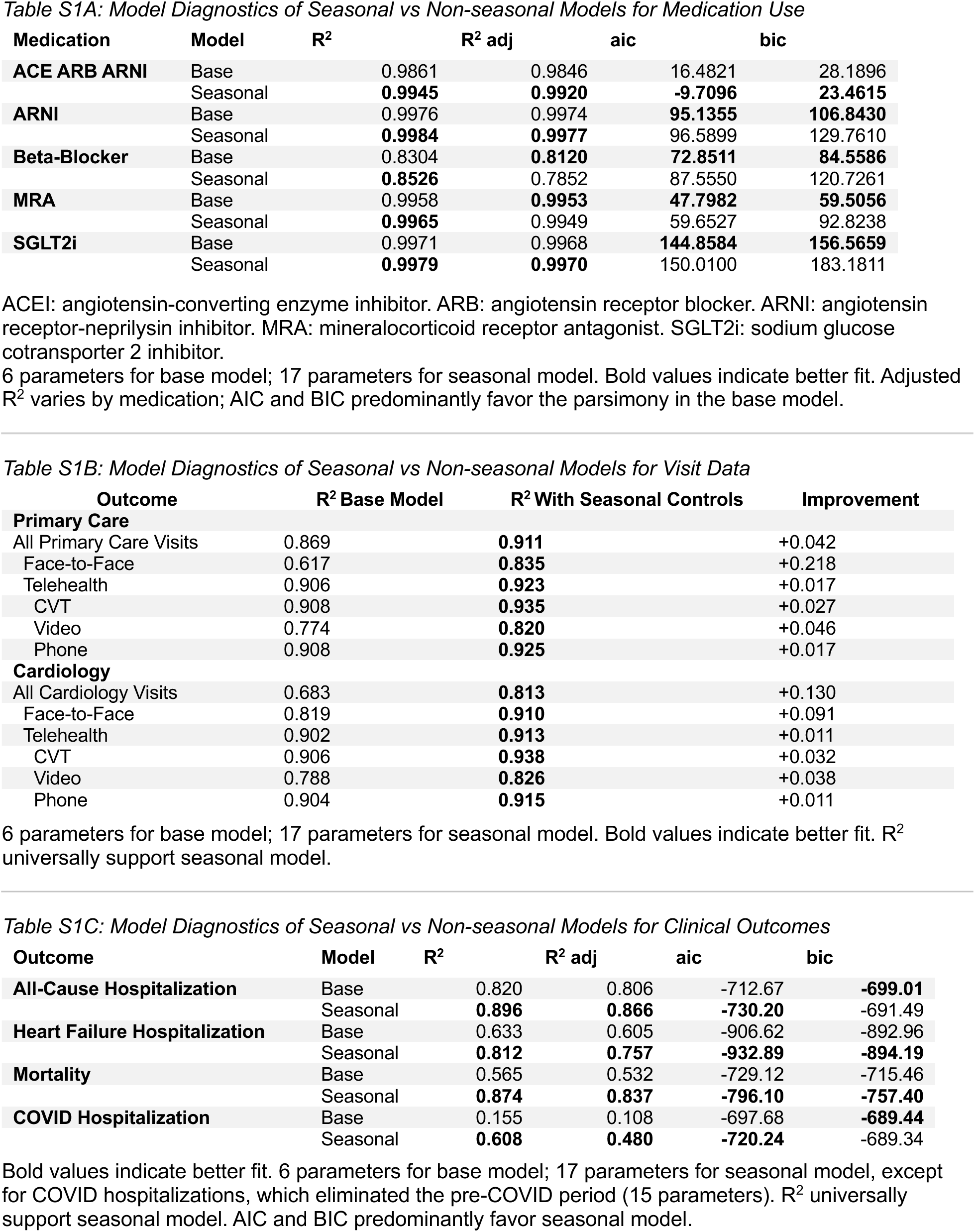
Sensitivity Analysis: Seasonal vs Non-seasonal Models for All Outcomes.

**Table S2:** Full Regression Results for Interrupted Time Series for Medication Utilization.

|  | ACE ARB ARNI | ARNI | Beta-blocker | MRA | SGLT2i |
| --- | --- | --- | --- | --- | --- |
| **Pre-COVID Trend (per period)** |  |  |  |  |  |
| Coeff. | 0.0085 | 0.40*** | 0.041*** | 0.14*** | 0.29*** |
| 95% CI | [-0.019, 0.036] | [0.38, 0.42] | [0.020, 0.062] | [0.13, 0.15] | [0.26, 0.33] |
| **Early COVID (Period 2)** |  |  |  |  |  |
| Level Change | -1.76* | -3.08** | 0.88** | -2.03** | -6.17*** |
| 95% CI | [-3.47, -0.062] | [-5.13, -1.04] | [0.34, 1.42] | [-3.33, -0.73] | [-9.00, -3.34] |
| Trend Change | 0.12* | 0.22** | -0.078*** | 0.14** | 0.48*** |
| 95% CI | [0.021, 0.22] | [0.089, 0.34] | [-0.12, -0.037] | [0.062, 0.22] | [0.30, 0.65] |
| **Late COVID (Period 3)** |  |  |  |  |  |
| Level Change | -1.77*** | -4.29** | -3.93*** | -7.18*** | -15.18*** |
| 95% CI | [-2.25, -1.28] | [-6.72, -1.85] | [-5.97, -1.90] | [-8.49, -5.87] | [-19.15, -11.21] |
| Trend Change | 0.14*** | 0.36*** | 0.093** | 0.33*** | 0.97*** |
| 95% CI | [0.12, 0.17] | [0.30, 0.43] | [0.040, 0.15] | [0.29, 0.36] | [0.86, 1.08] |

ACEI: angiotensin-converting enzyme inhibitor. ARB: angiotensin receptor blocker. ARNI: angiotensin receptor-neprilysin inhibitor. MRA: mineralocorticoid receptor antagonist. SGLT2i: sodium glucose cotransporter 2 inhibitor.
Presented results are from regression models with Newey-West robust standard errors to account for heteroskedasticity and autocorrelation as found in the time series data. Reference period: Pre-COVID (period 1).
\*p < 0.05, \*\*p < 0.01, \*\*\*p < 0.001 (based on confidence intervals not including zero). Level changes represent immediate shifts at period transitions; trend changes represent changes in the slope (rate of change per period).

**Table S3:** Full Regression Results for Interrupted Time Series for Primary Care Visits.

| Variable | All | Telehealth | Face-to-face | CVT | Video | Phone |
| --- | --- | --- | --- | --- | --- | --- |
| **Time Trends** |  |  |  |  |  |  |
| **Base Trend (monthly)** | -0.000<br>(0.000) | -0.000<br>(0.000) | -0.000<br>(0.000) | 0.000**<br>(0.000) | 0.000<br>(0.000) | -0.000<br>(0.000) |
| Period Effects (Level Changes) – ref. Pre COVID |  |  |  |  |  |  |
| **Early COVID** | 0.116***<br>(0.031) | 0.182***<br>(0.032) | -0.066***<br>(0.010) | -0.000*<br>(0.000) | -0.007***<br>(0.001) | 0.189***<br>(0.032) |
| **Late COVID** | 0.062***<br>(0.006) | 0.086***<br>(0.007) | -0.024***<br>(0.004) | -0.001***<br>(0.000) | 0.005***<br>(0.001) | 0.082***<br>(0.007) |
| Trend Changes (Period × Time Interactions) – ref. Pre-COVID x Time |  |  |  |  |  |  |
| **Early COVID × Time** | -0.002<br>(0.001) | -0.004**<br>(0.001) | 0.002***<br>(0.000) | -0.000<br>(0.000) | 0.000***<br>(0.000) | -0.005***<br>(0.001) |
| **Late COVID × Time** | -0.001**<br>(0.000) | -0.001***<br>(0.000) | 0.000<br>(0.000) | 0.000<br>(0.000) | -0.000<br>(0.000) | -0.001***<br>(0.000) |
| Seasonal Controls – ref. January |  |  |  |  |  |  |
| **February** | -0.017***<br>(0.003) | -0.011***<br>(0.002) | -0.005***<br>(0.001) | -0.000***<br>(0.000) | -0.000<br>(0.000) | -0.011***<br>(0.002) |
| **March** | 0.004<br>(0.011) | -0.002<br>(0.013) | 0.006*<br>(0.003) | 0.000<br>(0.000) | 0.000<br>(0.000) | -0.002<br>(0.013) |
| **April** | 0.005<br>(0.003) | 0.006<br>(0.006) | -0.001<br>(0.004) | -0.000<br>(0.000) | -0.000<br>(0.001) | 0.007<br>(0.006) |
| **May** | 0.001<br>(0.004) | 0.002<br>(0.004) | -0.001<br>(0.003) | -0.000<br>(0.000) | -0.001<br>(0.001) | 0.003<br>(0.004) |
| **June** | -0.002<br>(0.005) | -0.001<br>(0.006) | -0.001<br>(0.001) | -0.000<br>(0.000) | -0.001<br>(0.001) | 0.000<br>(0.006) |
| **July** | -0.002<br>(0.006) | -0.002<br>(0.005) | -0.001<br>(0.002) | -0.000<br>(0.000) | -0.001<br>(0.001) | -0.000<br>(0.005) |
| **August** | 0.009**<br>(0.003) | 0.005<br>(0.004) | 0.004**<br>(0.002) | 0.000<br>(0.000) | -0.001<br>(0.001) | 0.006<br>(0.004) |
| **September** | -0.000<br>(0.005) | -0.006<br>(0.004) | 0.006*<br>(0.003) | -0.000<br>(0.000) | -0.001<br>(0.001) | -0.006<br>(0.003) |
| **October** | 0.013*<br>(0.005) | -0.003<br>(0.004) | 0.016***<br>(0.003) | -0.000<br>(0.000) | -0.001<br>(0.001) | -0.002<br>(0.004) |
| **November** | -0.011*<br>(0.005) | -0.014*<br>(0.006) | 0.002<br>(0.002) | -0.000**<br>(0.000) | -0.001<br>(0.001) | -0.013*<br>(0.005) |
| **December** | -0.005<br>(0.004) | -0.005<br>(0.003) | -0.000<br>(0.002) | -0.000**<br>(0.000) | -0.001<br>(0.001) | -0.004<br>(0.004) |

Presented results are from regression models with Newey-West robust standard errors to account for heteroskedasticity and autocorrelation as found in the time series data. Reference period: Pre-COVID (period 1).
\*p < 0.05, \*\*p < 0.01, \*\*\*p < 0.001 (based on confidence intervals). Standard errors in parentheses. Level changes represent immediate shifts at period transitions; trend changes represent changes in the slope (rate of change per period).

**Table S4:** Full Regression Results of Interrupted Time Series for Cardiology Visits.

| Variable | All | Telehealth | Face-to-face | CVT | Video | Phone |
| --- | --- | --- | --- | --- | --- | --- |
| **Time Trends** |  |  |  |  |  |  |
| **Base Trend (monthly)** | -0.000***<br>(0.000) | 0.000<br>(0.000) | -0.000**<br>(0.000) | 0.000<br>(0.000) | 0.000<br>(0.000) | -0.000<br>(0.000) |
| Period Effects (Level Changes) – ref. Pre COVID |  |  |  |  |  |  |
| **Early COVID** | 0.032**<br>(0.011) | 0.086***<br>(0.013) | -0.054***<br>(0.006) | -0.001***<br>(0.000) | -0.003***<br>(0.000) | 0.090***<br>(0.013) |
| **Late COVID** | -0.007*<br>(0.003) | 0.013***<br>(0.002) | -0.020***<br>(0.002) | -0.001***<br>(0.000) | 0.002***<br>(0.000) | 0.012***<br>(0.002) |
| Trend Changes (Period × Time Interactions) – ref. Pre-COVID x Time |  |  |  |  |  |  |
| **Early COVID × Time** | -0.000<br>(0.000) | -0.002***<br>(0.001) | 0.002***<br>(0.000) | 0.000**<br>(0.000) | 0.000***<br>(0.000) | -0.002***<br>(0.001) |
| **Late COVID × Time** | 0.001***<br>(0.000) | -0.000<br>(0.000) | 0.001***<br>(0.000) | 0.000***<br>(0.000) | -0.000<br>(0.000) | -0.000<br>(0.000) |
| Seasonal Controls (ref. January) |  |  |  |  |  |  |
| **February** | -0.004***<br>(0.001) | -0.001***<br>(0.000) | -0.003**<br>(0.001) | -0.000<br>(0.000) | -0.000<br>(0.000) | -0.001***<br>(0.000) |
| **March** | 0.003<br>(0.004) | -0.004<br>(0.005) | 0.007***<br>(0.002) | 0.000<br>(0.000) | -0.000<br>(0.000) | -0.004<br>(0.005) |
| **April** | 0.004**<br>(0.001) | 0.002<br>(0.003) | 0.002<br>(0.002) | 0.000<br>(0.000) | -0.000<br>(0.000) | 0.002<br>(0.003) |
| **May** | 0.003<br>(0.002) | 0.001<br>(0.002) | 0.002<br>(0.002) | 0.000*<br>(0.000) | -0.000<br>(0.000) | 0.001<br>(0.002) |
| **June** | 0.001<br>(0.002) | -0.000<br>(0.001) | 0.001<br>(0.001) | 0.000*<br>(0.000) | -0.000<br>(0.000) | -0.000<br>(0.001) |
| **July** | -0.001<br>(0.002) | -0.001<br>(0.001) | 0.000<br>(0.001) | -0.000<br>(0.000) | -0.000<br>(0.000) | -0.001<br>(0.001) |
| **August** | 0.004**<br>(0.001) | 0.000<br>(0.001) | 0.004***<br>(0.001) | 0.000**<br>(0.000) | -0.000<br>(0.000) | 0.000<br>(0.001) |
| **September** | 0.000<br>(0.002) | -0.002<br>(0.001) | 0.002<br>(0.001) | 0.000<br>(0.000) | -0.000<br>(0.000) | -0.001<br>(0.001) |
| **October** | 0.002<br>(0.001) | -0.001<br>(0.001) | 0.003*<br>(0.001) | 0.000**<br>(0.000) | -0.000<br>(0.000) | -0.001<br>(0.001) |
| **November** | -0.005**<br>(0.002) | -0.003*<br>(0.002) | -0.002<br>(0.001) | 0.000<br>(0.000) | -0.000<br>(0.000) | -0.003*<br>(0.001) |
| **December** | -0.004*<br>(0.002) | -0.002***<br>(0.001) | -0.002<br>(0.002) | -0.000<br>(0.000) | -0.000<br>(0.000) | -0.002**<br>(0.001) |

Presented results are from regression models with Newey-West robust standard errors to account for heteroskedasticity and autocorrelation as found in the time series data. Reference period: Pre-COVID (period 1).
\*p < 0.05, \*\*p < 0.01, \*\*\*p < 0.001 (based on confidence intervals). Standard errors in parentheses. Level changes represent immediate shifts at period transitions; trend changes represent changes in the slope (rate of change per period).

**Table S5:** Full Regression Results of Interrupted Time Series for Hospitalization and Mortality.

| Variable | All-cause Hospitalizations | Heart Failure Hospitalizations | Mortality | COVID-19 Hospitalizations |
| --- | --- | --- | --- | --- |
| Base Trend (monthly) | -0.0001<br>(0.0001) | -0.0000<br>(0.0000) | 0.0001<br>(0.0001) | -0.0000<br>(0.0000) |
| Period Effects (Level Changes) – ref. Pre COVID |  |  |  |  |
| Early COVID | -0.0190***<br>(0.0026) | -0.0031***<br>(0.0005) | 0.0024*<br>(0.0011) | 0.0000<br>(.) |
| Late COVID | -0.0094***<br>(0.0008) | -0.0015***<br>(0.0002) | 0.0056***<br>(0.0011) | 0.0007<br>(0.0004) |
| Trend Changes (Period × Time Interactions) – ref. Pre-COVID x Time |  |  |  |  |
| Early COVID × Time | 0.0005***<br>(0.0001) | 0.0001***<br>(0.0000) | -0.0000<br>(0.0001) | 0.0000<br>(.) |
| Late COVID × Time | 0.0001<br>(0.0001) | 0.0000<br>(0.0000) | -0.0001<br>(0.0001) | -0.0000<br>(0.0000) |
| Monthly Effects |  |  |  |  |
| **February** | -0.0026***<br>(0.0006) | -0.0004*<br>(0.0002) | -0.0026***<br>(0.0005) | -0.0012*<br>(0.0006) |
| **March** | 0.0015<br>(0.0008) | 0.0003<br>(0.0002) | -0.0023**<br>(0.0008) | -0.0016*<br>(0.0006) |
| **April** | -0.0008<br>(0.0013) | -0.0003<br>(0.0003) | -0.0032***<br>(0.0008) | -0.0015*<br>(0.0006) |
| **May** | 0.0003<br>(0.0007) | -0.0002<br>(0.0002) | -0.0033***<br>(0.0008) | -0.0015*<br>(0.0006) |
| **June** | -0.0005<br>(0.0007) | -0.0005*<br>(0.0002) | -0.0043***<br>(0.0007) | -0.0015*<br>(0.0006) |
| **July** | -0.0007<br>(0.0008) | -0.0007***<br>(0.0002) | -0.0033***<br>(0.0007) | -0.0012*<br>(0.0006) |
| **August** | 0.0001<br>(0.0006) | -0.0005*<br>(0.0002) | -0.0036***<br>(0.0007) | -0.0010<br>(0.0006) |
| **September** | -0.0014<br>(0.0008) | -0.0008***<br>(0.0002) | -0.0038***<br>(0.0007) | -0.0011<br>(0.0006) |
| **October** | -0.0001<br>(0.0008) | -0.0002<br>(0.0003) | -0.0029***<br>(0.0006) | -0.0012*<br>(0.0006) |
| **November** | -0.0020**<br>(0.0006) | -0.0005*<br>(0.0002) | -0.0027***<br>(0.0006) | -0.0010<br>(0.0006) |
| **December** | -0.0004<br>(0.0006) | -0.0002<br>(0.0002) | -0.0007<br>(0.0006) | -0.0006<br>(0.0007) |

Presented results are from regression models with Newey-West robust standard errors to account for heteroskedasticity and autocorrelation as found in the time series data. Reference period: Pre-COVID (period 1).
\*p < 0.05, \*\*p < 0.01, \*\*\*p < 0.001 (based on confidence intervals not including zero). Level changes represent immediate shifts at period transitions; trend changes represent changes in the slope (rate of change per period).

**Table S6:** Demographics and Comorbidities for Imputed Datasets and Complete Cases.

| Characteristic | Multiple Imputation (Primary) | Complete Case (Sensitivity) |
| --- | --- | --- |
| Sample Size | N = 210535 | N = 139169 |
| **DEMOGRAPHICS** |  |  |
| Age, years | 73.1 ± 10.6 | 72.5 ± 10.2 |
| Female sex | 4639 (2.2%) | 3135 (2.3%) |
| Race/Ethnicity |  |  |
| White | 146221 (69.5%) | 97082 (69.8%) |
| Black | 40172 (19.1%) | 27724 (19.9%) |
| Hispanic | 9143 (4.3%) | 4899 (3.5%) |
| Other | 14999 (7.1%) | 9464 (6.8%) |
| **VITAL SIGNS AND LABS** |  |  |
| BMI, kg/m<sup>2</sup> | 29.8 ± 6.7 | 30.0 ± 6.7 |
| Heart rate, bpm | 75.0 ± 14.4 | 75.1 ± 14.5 |
| Systolic BP, mmHg | 126.5 ± 19.4 | 126.6 ± 19.5 |
| Diastolic BP, mmHg | 72.3 ± 11.5 | 72.4 ± 11.5 |
| Hemoglobin, g/dL | 13.0 ± 2.2 | 13.0 ± 2.2 |
| eGFR, mL/min/1.73m<sup>2</sup> | 63.0 ± 27.9 | 62.3 ± 27.8 |
| Ejection Fraction, % | 32.5 ± 14.5 | 32.7 ± 14.6 |
| **COMORBIDITIES** |  |  |
| Atrial fibrillation | 91798 (43.6%) | 63482 (45.6%) |
| Coronary artery disease | 151031 (71.7%) | 103649 (74.5%) |
| Chronic kidney disease | 111959 (53.2%) | 78141 (56.1%) |
| COPD | 18067 (8.6%) | 12892 (9.3%) |
| Diabetes mellitus | 129413 (61.5%) | 88702 (63.7%) |
| Hyperlipidemia | 161487 (76.7%) | 112401 (80.8%) |
| Hypertension | 187912 (89.3%) | 127624 (91.7%) |
| Cerebrovascular accident | 31063 (14.8%) | 21831 (15.7%) |
| Alcohol use disorder | 25552 (12.1%) | 18158 (13.0%) |
| Substance use disorder | 18328 (8.7%) | 13198 (9.5%) |
| Smoking | 39741 (18.9%) | 28020 (20.1%) |
| Depression | 54828 (26.0%) | 38411 (27.6%) |
| Peripheral artery disease | 107021 (50.8%) | 74716 (53.7%) |
| Cancer | 37158 (17.6%) | 25821 (18.6%) |
| **OUTCOMES** |  |  |
| Death | 92459 (43.9%) | 68172 (49.0%) |
| First hospitalization | 61380 (29.2%) | 49630 (35.7%) |
| Composite endpoint | 119562 (56.8%) | 89602 (64.4%) |

BMI, body mass index; BP, blood pressure; COPD, chronic obstructive pulmonary disease; eGFR, estimated glomerular filtration rate.
Baseline characteristics under the Multiple Imputation header are presented for the first imputed dataset (m=1), representative of all imputed datasets. Complete cases included patients with valid survival times, event status, and no missing values in covariates after survival data preparation (n=139,169; 66.1%). Baseline characteristics imputed closest to cohort entry, which may not reflect patients' clinical status throughout the 6 years of follow up, particularly given the COVID-related disruptions to the healthcare system.

**Table S7:** Sensitivity Analysis: Cox Proportional Hazard Models for Mortality and Hospitalization.

| Variable | All-Cause Mortality<br>HR (95% CI) | All-Cause Hospitalization<br>HR (95% CI) | Composite Endpoint<br>HR (95% CI) |
| --- | --- | --- | --- |
| **Demographics** |  |  |  |
| Age at entry (per year) | 1.034 (1.033-1.035)*** | 0.997 (0.996-0.999)*** | 1.015 (1.014-1.016)*** |
| Female sex | 0.804 (0.757-0.854)*** | 0.907 (0.853-0.965)** | 0.885 (0.843-0.929)*** |
| **Race/Ethnicity** |  |  |  |
| White (reference) | 1 | 1 | 1 |
| Black | 0.964 (0.944-0.984)** | 1.371 (1.340-1.402)*** | 1.148 (1.127-1.168)*** |
| Hispanic | 0.900 (0.862-0.939)*** | 1.168 (1.115-1.224)*** | 1.021 (0.984-1.058) |
| Other | 0.933 (0.904-0.962)*** | 0.940 (0.906-0.976)** | 0.955 (0.929-0.981)** |
| **Geographic Region** |  |  |  |
| Midwest (reference) | 1 | 1 | 1 |
| Northeast | 0.981 (0.955-1.006) | 0.978 (0.948-1.009) | 0.984 (0.962-1.007) |
| South | 1.017 (0.998-1.037) | 0.971 (0.950-0.993)* | 0.989 (0.972-1.005) |
| West | 1.042 (1.018-1.066)** | 0.978 (0.951-1.006) | 1.007 (0.987-1.028) |
| **Clinical Parameters** |  |  |  |
| BMI (per kg/m<sup>2</sup>) | 0.991 (0.990-0.992)*** | 1.000 (0.998-1.001) | 0.998 (0.996-0.999)*** |
| Pulse (per bpm) | 1.001 (1.000-1.001)* | 0.998 (0.998-0.999)*** | 0.999 (0.999-1.000)** |
| Systolic BP (per mmHg) | 1.003 (1.003-1.004)*** | 1.003 (1.003-1.004)*** | 1.004 (1.003-1.004)*** |
| Diastolic BP (per mmHg) | 0.995 (0.994-0.996)*** | 1.000 (0.999-1.001) | 0.998 (0.997-0.998)*** |
| Hemoglobin (per g/dL) | 0.978 (0.975-0.982)*** | 1.009 (1.005-1.013)*** | 1.000 (0.997-1.003) |
| eGFR (per mL/min/1.73m<sup>2</sup>) | 0.997 (0.996-0.997)*** | 1.000 (1.000-1.000)* | 0.999 (0.998-0.999)*** |
| LVEF | 0.997 (0.997-0.998)*** | 0.984 (0.984-0.985)*** | 0.992 (0.992-0.993)*** |
| **Comorbidities** |  |  |  |
| Atrial fibrillation | 1.082 (1.065-1.099)*** | 1.095 (1.075-1.115)*** | 1.078 (1.064-1.093)*** |
| Coronary artery disease | 1.057 (1.037-1.077)*** | 1.208 (1.182-1.235)*** | 1.098 (1.081-1.116)*** |
| Chronic kidney disease | 1.130 (1.106-1.155)*** | 1.150 (1.120-1.180)*** | 1.126 (1.105-1.148)*** |
| COPD | 1.153 (1.125-1.182)*** | 1.067 (1.035-1.100)*** | 1.097 (1.072-1.121)*** |
| Diabetes mellitus | 1.050 (1.028-1.072)*** | 1.009 (0.985-1.034) | 1.031 (1.013-1.050)** |
| Hyperlipidemia | 0.924 (0.906-0.943)*** | 1.024 (0.999-1.049) | 0.957 (0.940-0.974)*** |
| Hypertension | 0.994 (0.964-1.025) | 1.180 (1.137-1.225)*** | 1.070 (1.042-1.098)*** |
| Cerebrovascular disease | 1.098 (1.077-1.121)*** | 1.090 (1.065-1.117)*** | 1.088 (1.069-1.107)*** |
| Alcohol use disorder | 1.115 (1.087-1.144)*** | 1.128 (1.097-1.160)*** | 1.116 (1.092-1.140)*** |
| Substance use disorder | 1.135 (1.101-1.169)*** | 1.230 (1.193-1.269)*** | 1.198 (1.168-1.228)*** |
| Smoking | 1.213 (1.188-1.238)*** | 1.151 (1.125-1.178)*** | 1.164 (1.144-1.185)*** |
| Depression | 1.056 (1.037-1.075)*** | 1.140 (1.118-1.163)*** | 1.103 (1.087-1.120)*** |
| Peripheral artery disease | 1.168 (1.146-1.190)*** | 1.153 (1.127-1.179)*** | 1.135 (1.117-1.154)*** |
| Cancer | 1.052 (1.032-1.072)*** | 1.106 (1.081-1.132)*** | 1.043 (1.026-1.061)*** |
| **Model Statistics** |  |  |  |
| C-statistic | 0.643 | 0.618 | 0.595 |

BMI, body mass index; BP, blood pressure; COPD, chronic obstructive pulmonary disease; eGFR, estimated glomerular filtration rate; HR, hazard ratio; LVEF, left ventricular ejection fraction.
Statistical significance: \*P < 0.05, \*\*P < 0.01, \*\*\*P < 0.001
Note: HR results from complete case analysis (N = 139,169); C statistic from same dataset. Models adjusted for all variables shown. Composite endpoint defined as time to first occurrence of all-cause mortality or heart failure hospitalization. White race serves as the reference category for race/ethnicity comparisons; Midwest serves as the reference category for geographic comparisons.

**Table S8:**
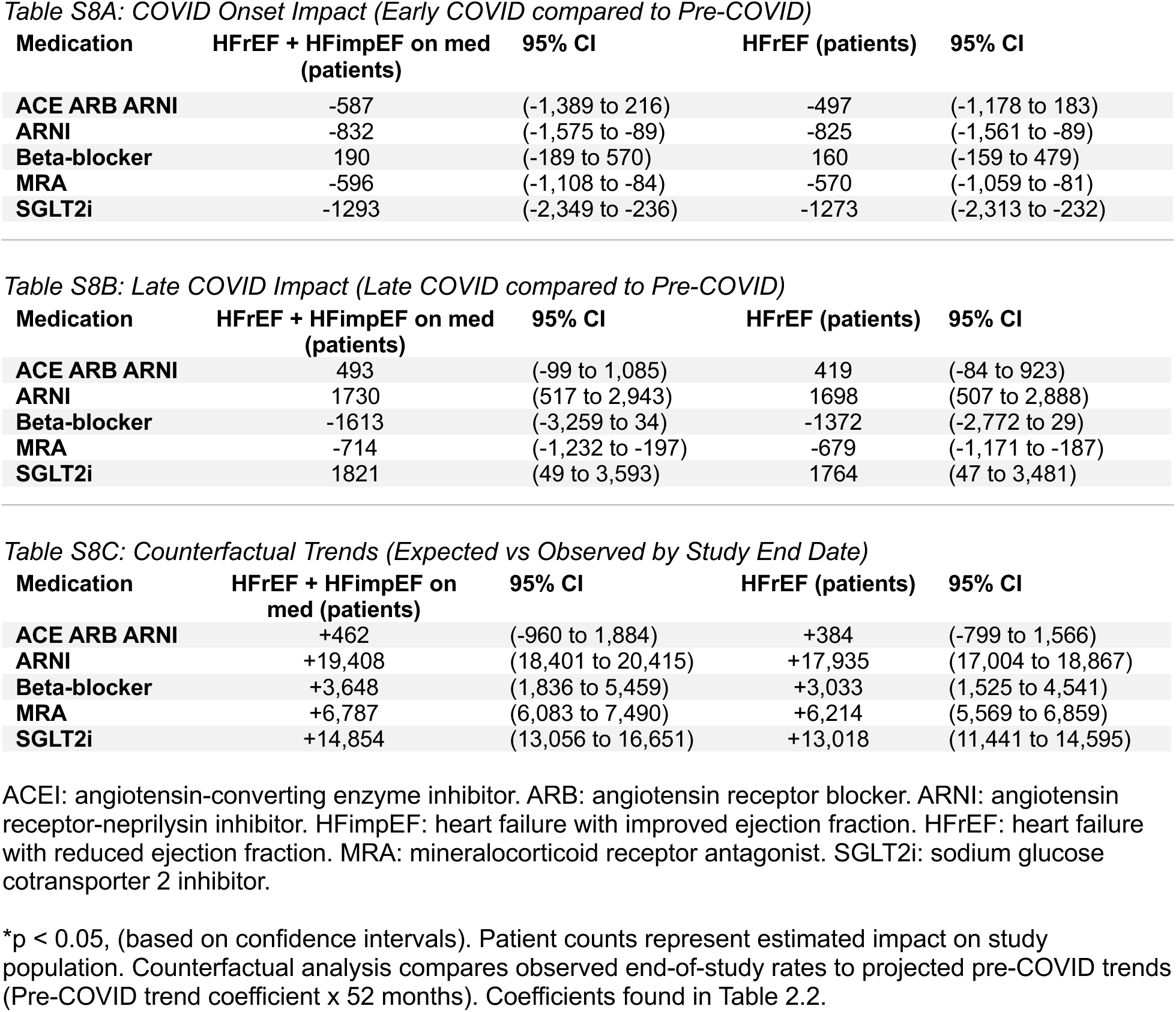
Estimated Number of Patients for GDMT Measures.

**Table S9:**
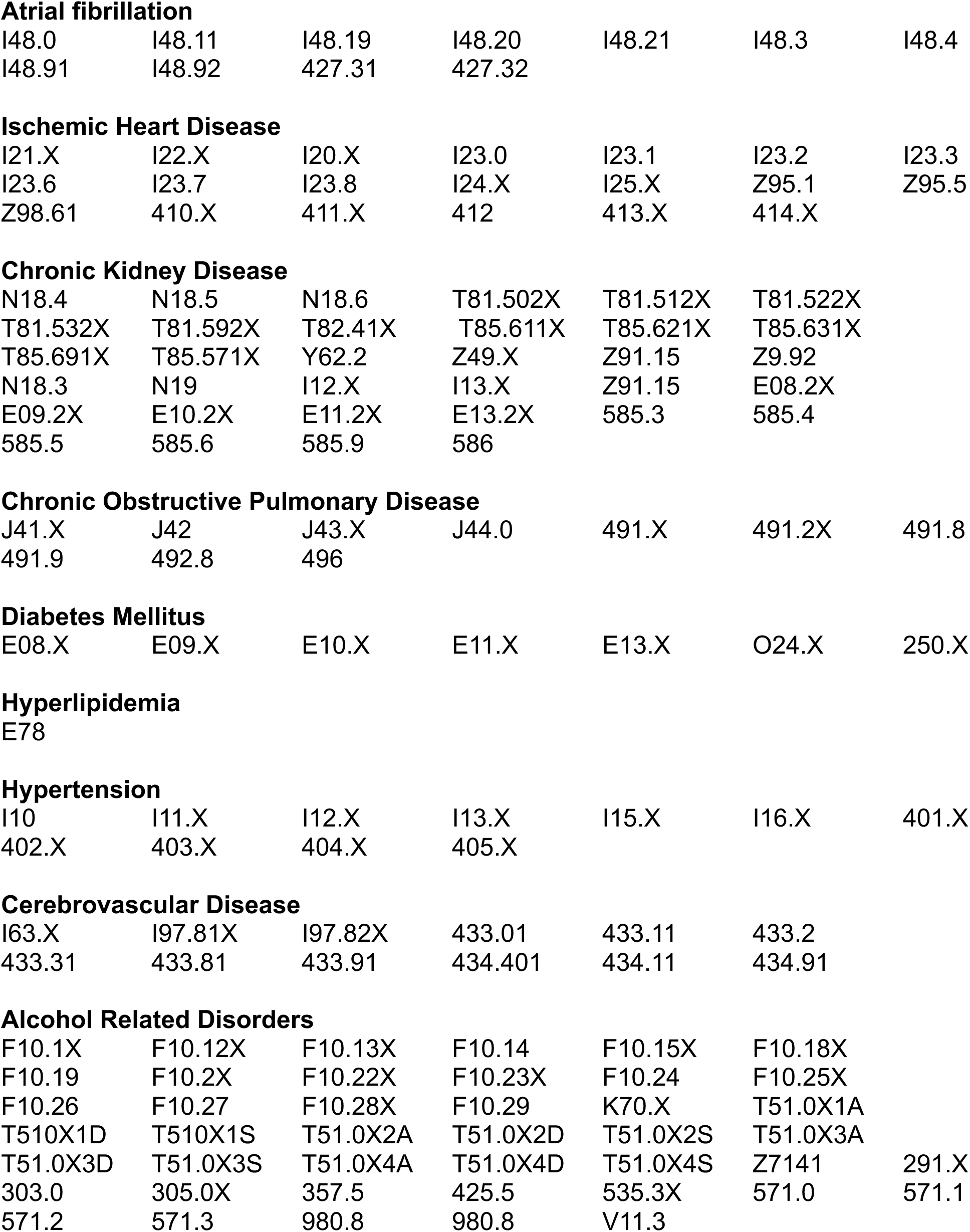
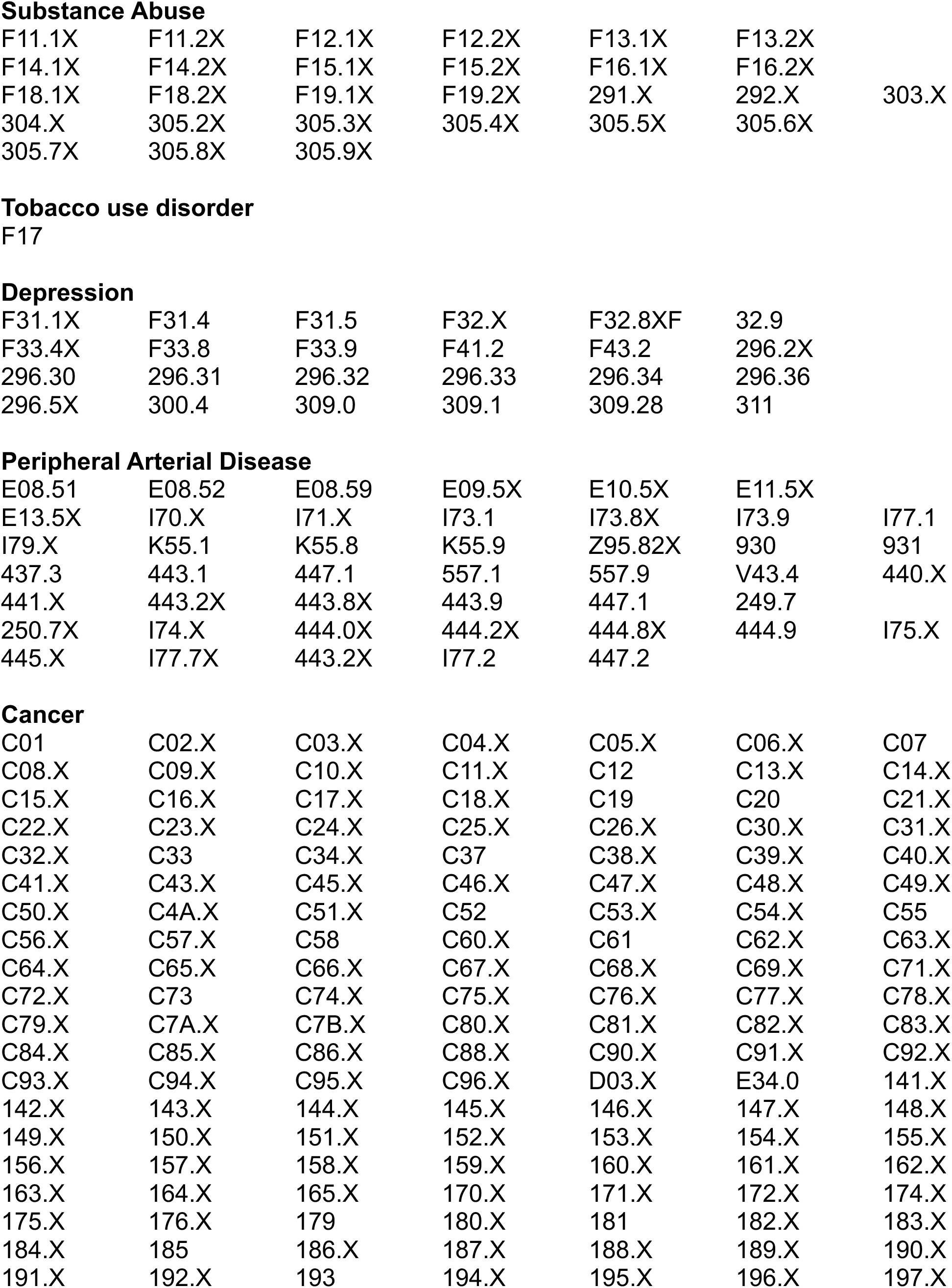
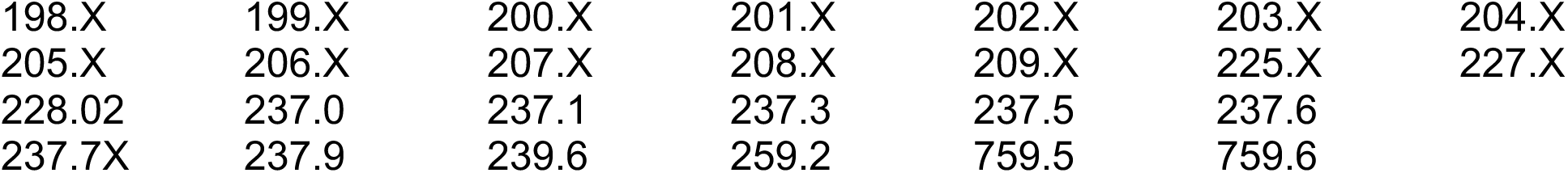
ICD Codes.

**Figure S1:**
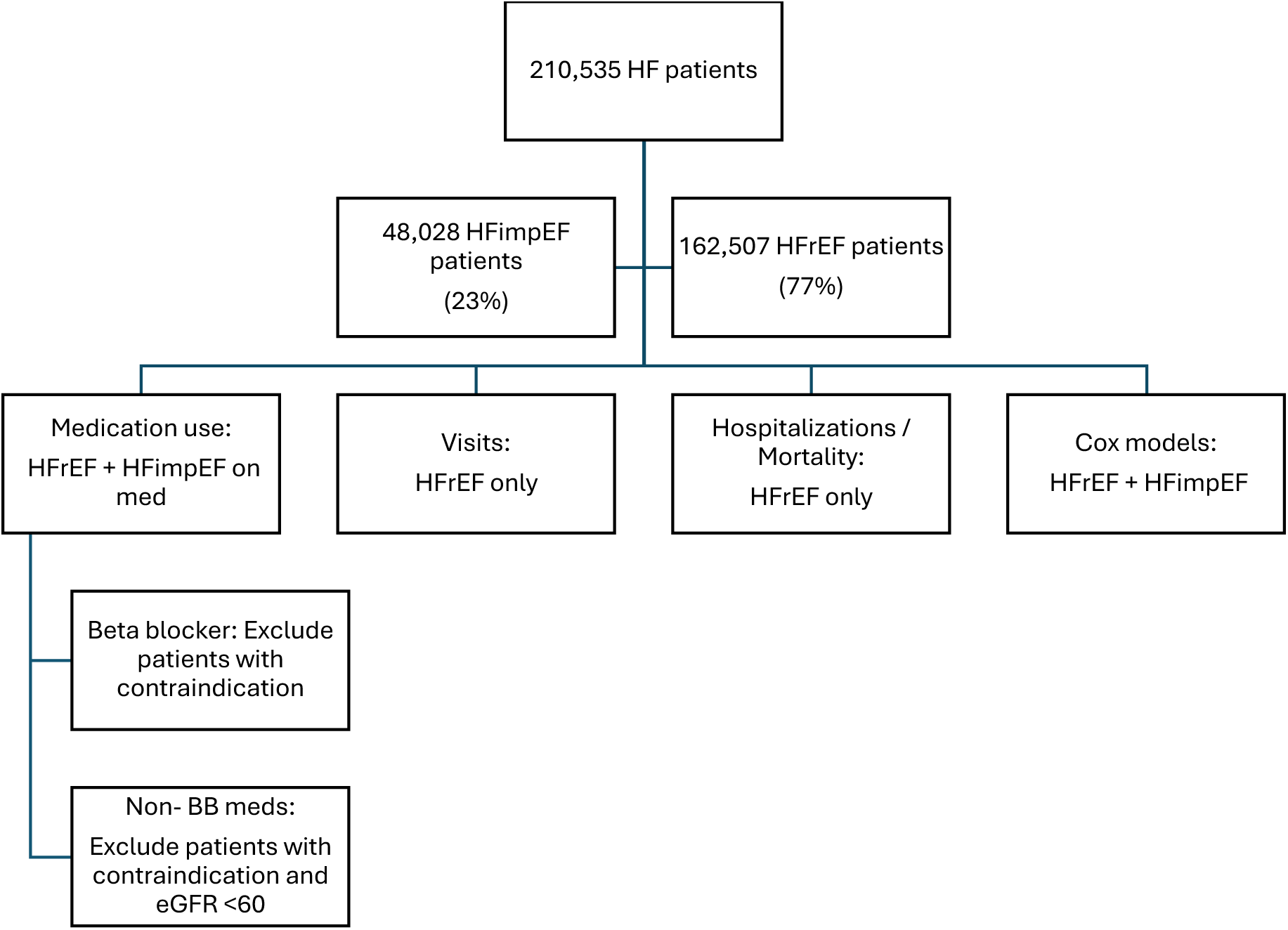
Selection of final cohort for analyses BB: Beta blocker. eGFR: estimated glomerular filtration rate. HF: heart failure. HFimpEF: heart failure with improved ejection fraction. HFrEF: heart failure with reduced ejection fraction.

**Figure S2:**
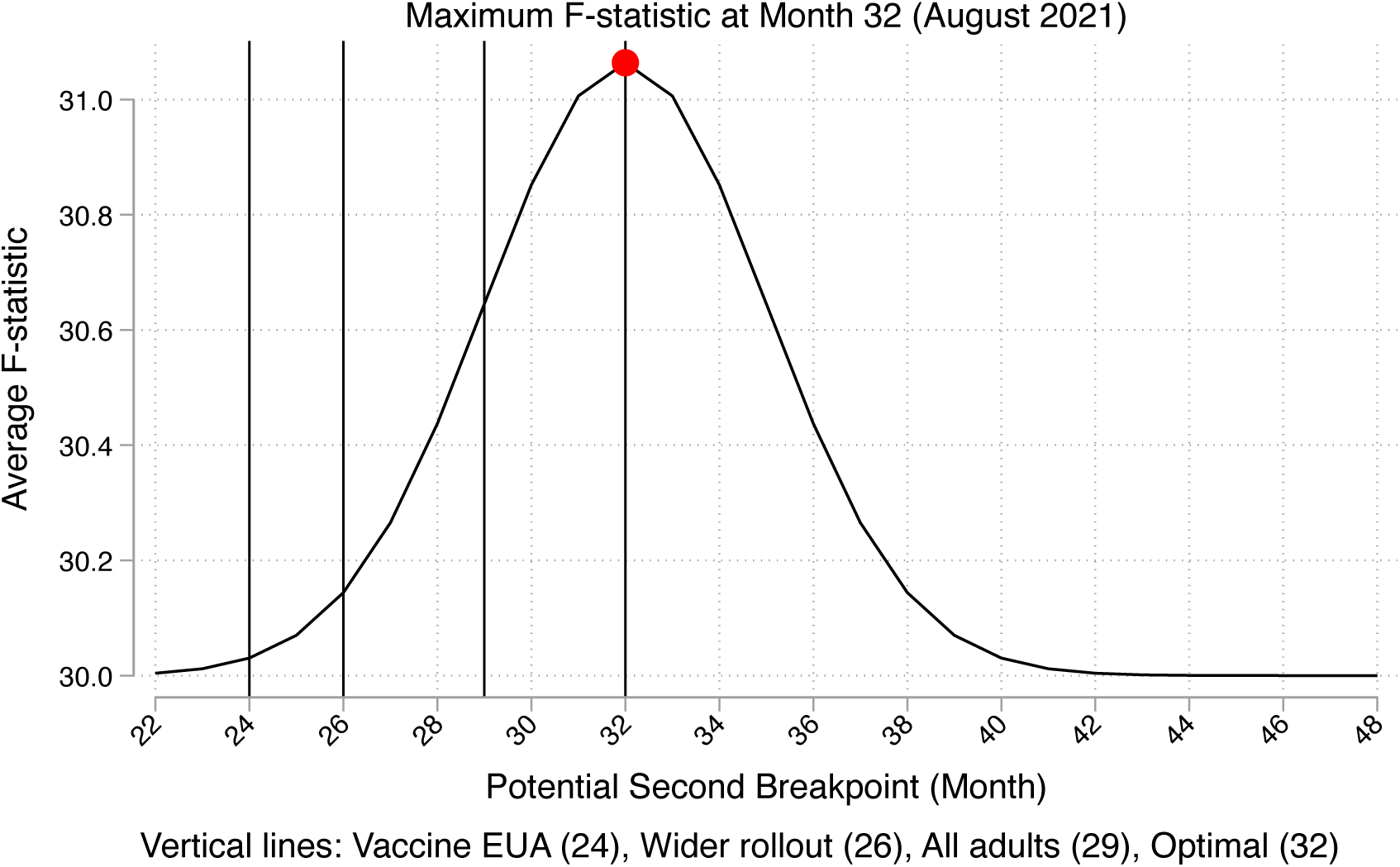
Optimal Breakpoint Selection Based on Variance Weighted Averages of Combined F-statistic for Hospitalizations and Mortality March 2020 marked the declaration of a National Pandemic Emergency in the United States so was chosen as breakpoint 1.^20^ Optimal breakpoint 2 was determined using structural break detection methods, testing all potential breakpoints from month 22 to 48 and noting August 2021 as the superior model fit across outcomes;^21^ specifically, systematic F-statistic optimization using variance weighted averages noted the highest combined F-statistic at month 32. Clinically this corresponded to the peak of the Delta variant, with the highest COVID hospitalizations since the initial surge as well as increased vaccine availability and eligibility for the general population.^22^

**Figure S3:**
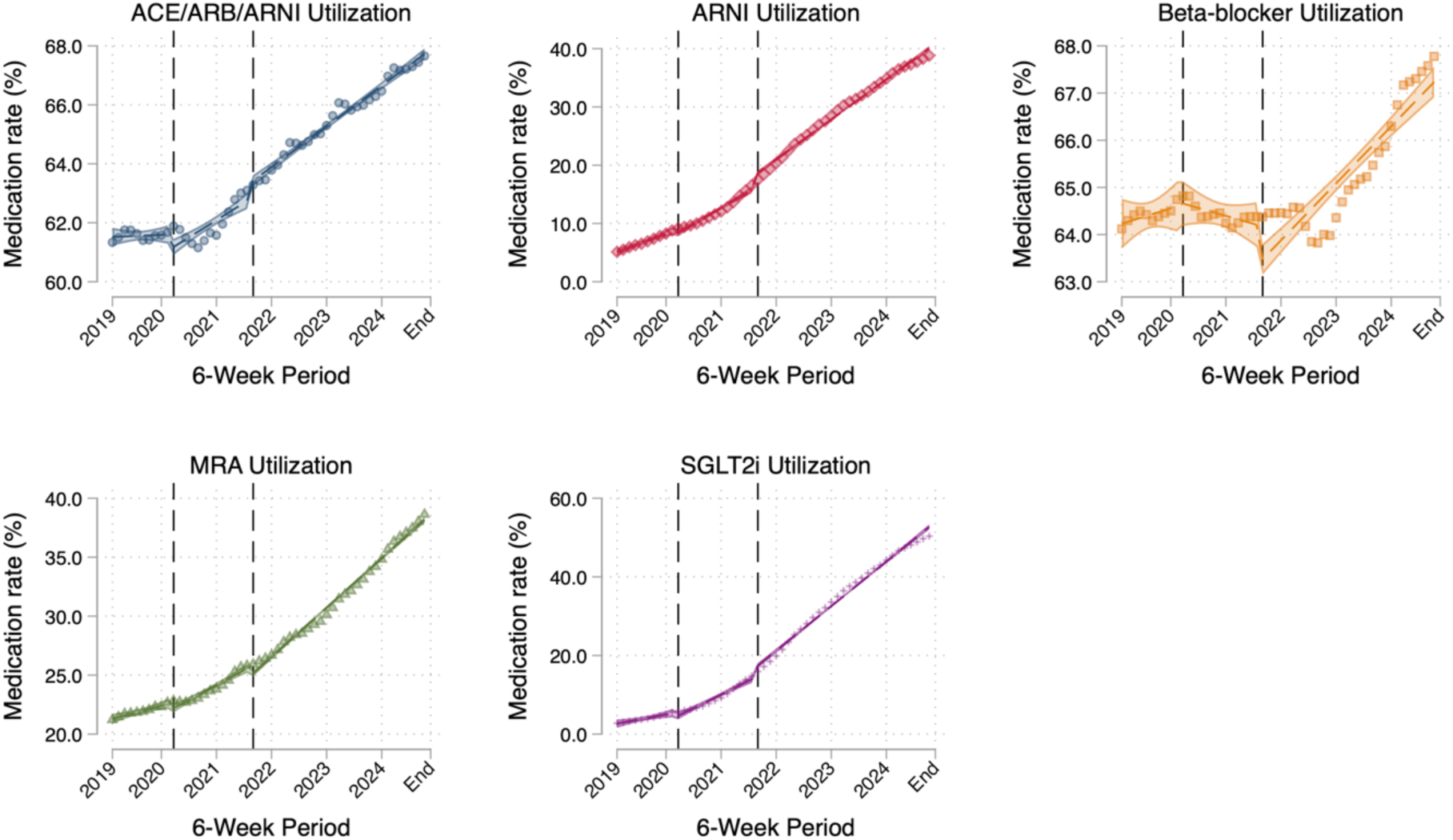
Individual Heart Failure Medication Utilization Trends Between 2019 and 2024 ACEI: angiotensin-converting enzyme inhibitor. ARB: angiotensin receptor blocker. ARNI: angiotensin receptor-neprilysin inhibitor. MRA: mineralocorticoid receptor antagonist. SGLT2i: sodium glucose cotransporter 2 inhibitor. First vertical line indicates transition from pre-COVID to early COVID phase (March 2020). Second vertical line indicates transition from early COVID to late COVID phase (August 2021).

**Figure S4:**
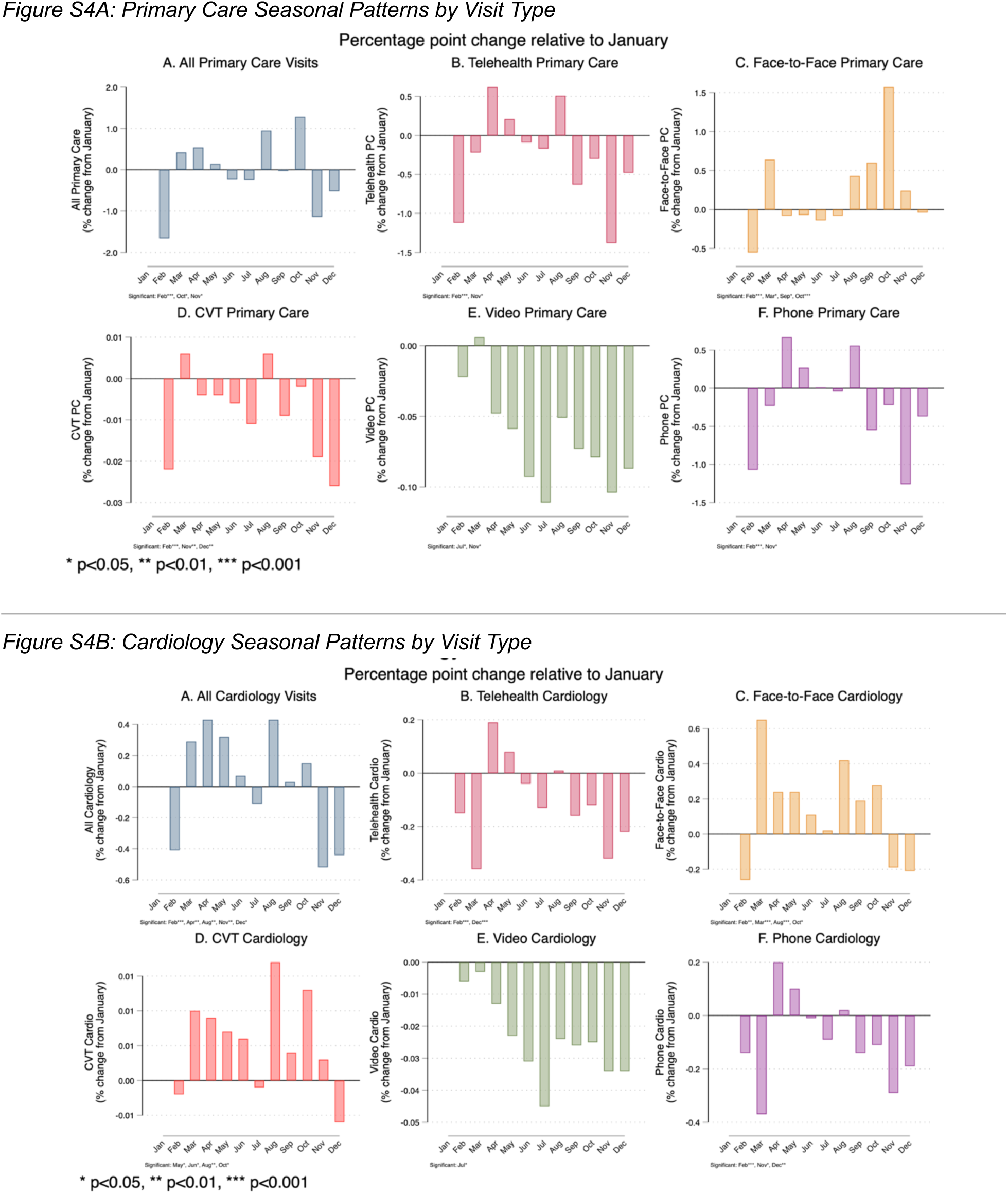
Seasonal Effects on Primary Care and Cardiology Visits.

**Figure S5:**
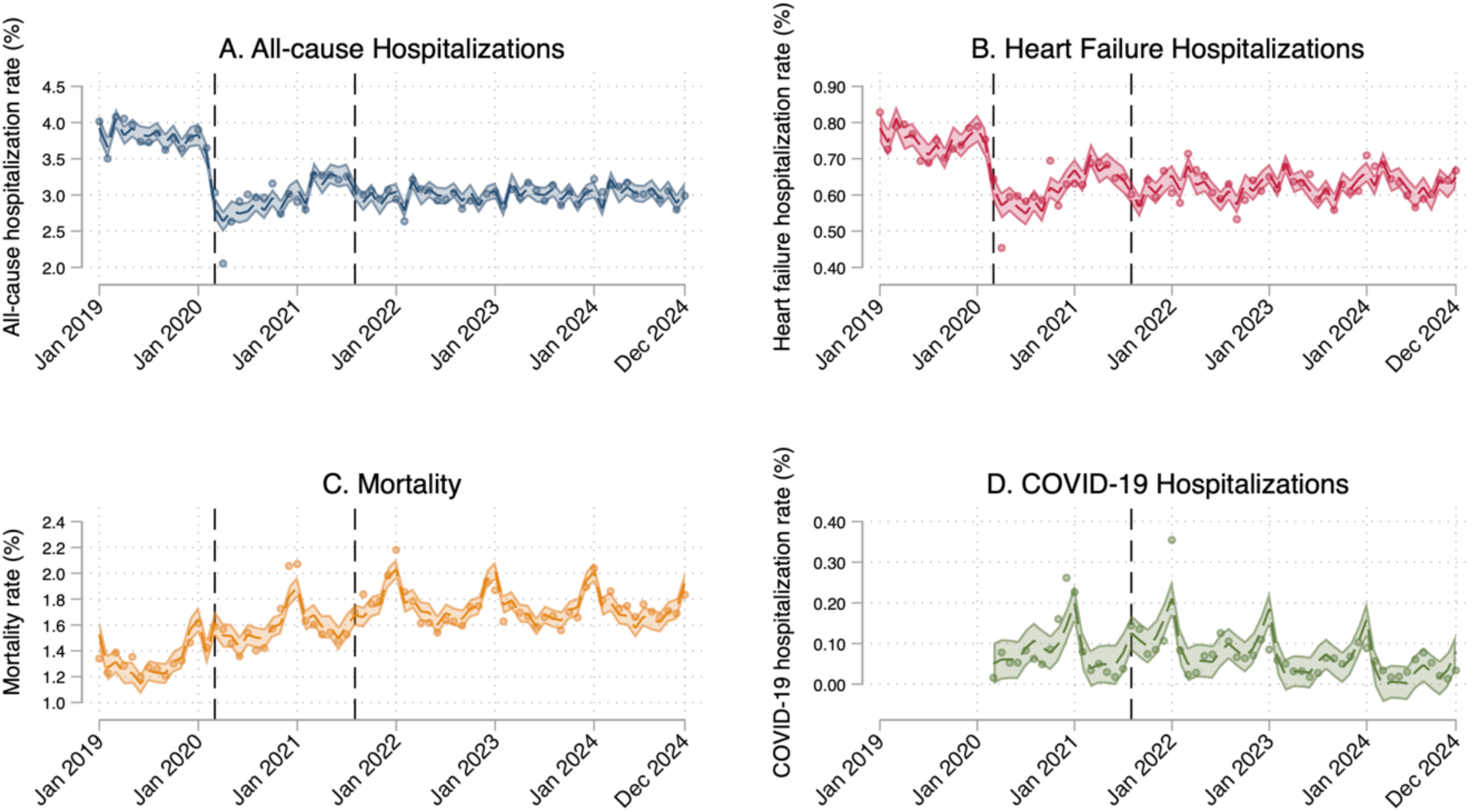
Individual Hospitalization and Mortality Trends Between 2019 and 2024 First vertical line indicates transition from pre-COVID to early COVID phase (March 2020). Second vertical line indicates transition from early COVID to late COVID phase (August 2021). Shaded areas represent 95% confidence intervals.

**Figure S6:**
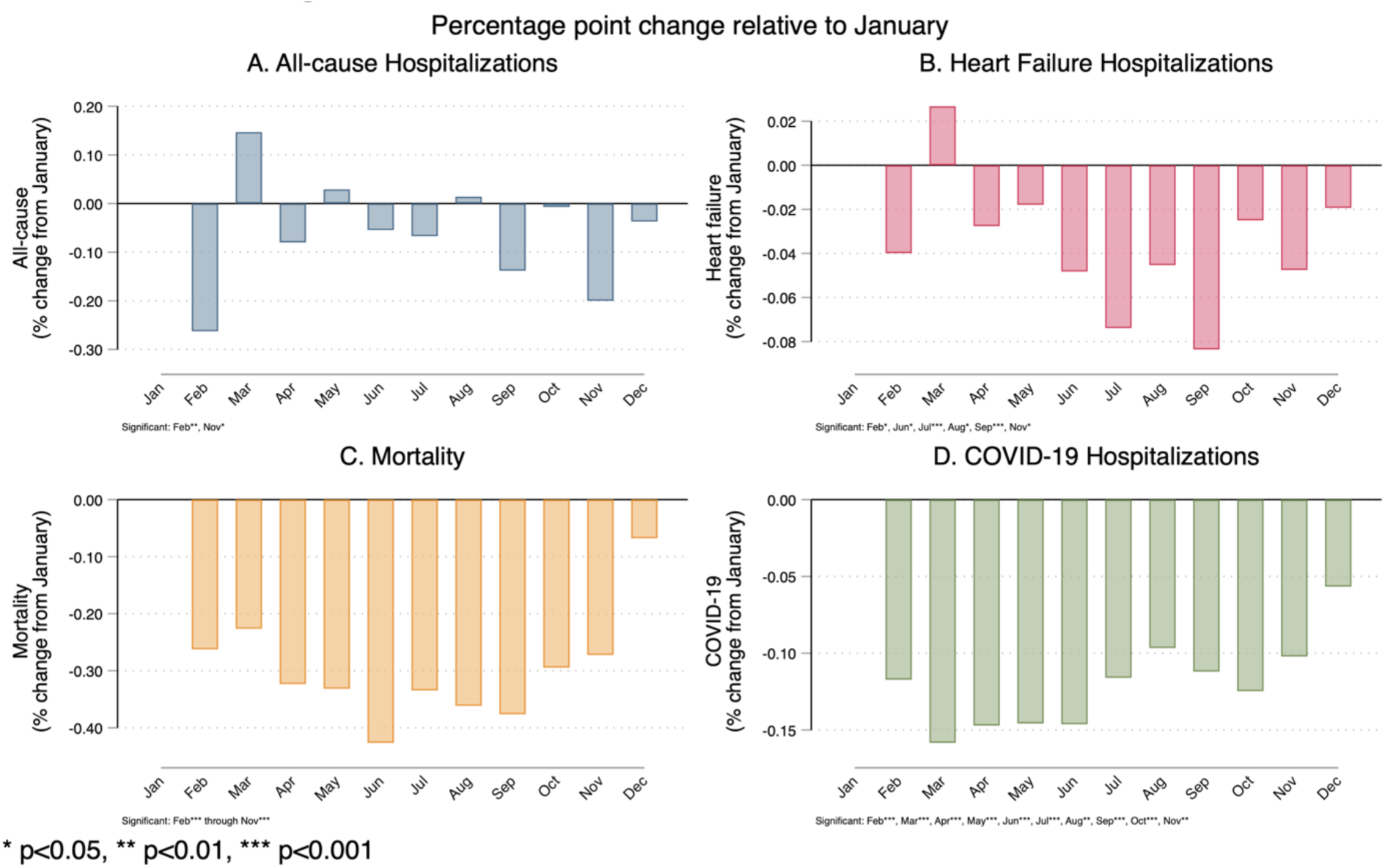
Seasonal Effects on Hospitalization and Mortality.

**Figure S7:**
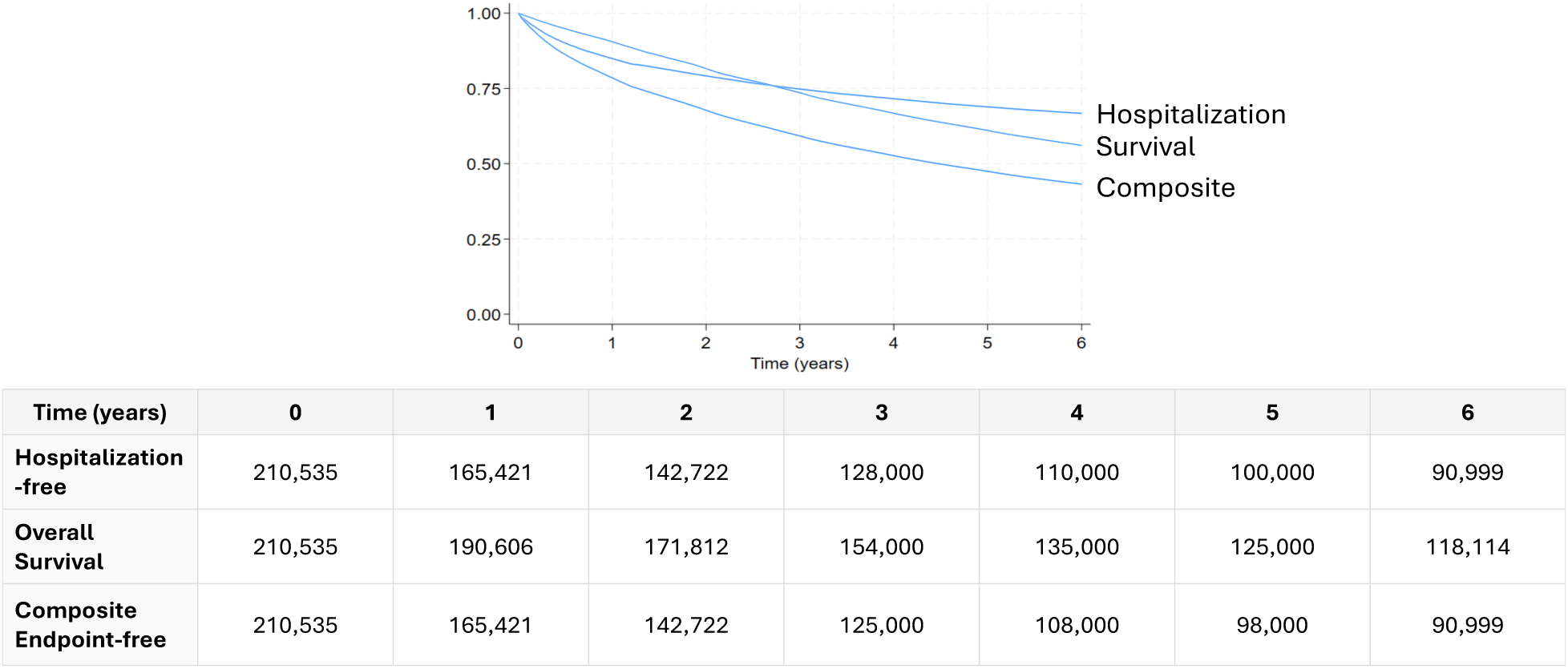
Kaplan Meier Curves for Time-To-Event Outcomes.

